# Transport-based transfer learning on Electronic Health Records: Application to detection of treatment disparities

**DOI:** 10.1101/2024.03.27.24304781

**Authors:** Wanxin Li, Saad Ahmed, Yongjin P. Park, Khanh Dao Duc

## Abstract

Electronic Health Records (EHRs) sampled from different populations can introduce unwanted biases, limit individual-level data sharing, and make the data and fitted model hardly transferable across different population groups. In this context, our main goal is to design an effective method to transfer knowledge between population groups, with computable guarantees for suitability, and that can be applied to quantify treatment disparities. For a model trained in an embedded feature space of one subgroup, our proposed framework Optimal Transport-based Transfer Learning for EHRs (*OTTEHR*) combines feature embedding of the data and unbalanced optimal transport (OT) for domain adaptation to another population group. To test our method, we processed and divided the MIMIC-III and MIMIC-IV databases into multiple population groups using ICD codes and multiple labels. We derive a theoretical bound for the generalization error of our method, and interpret it in terms of the Wasserstein distance, unbalancedness between the source and target domains, and labeling divergence, which can be used as a guide for assessing the suitability of binary classification and regression tasks. In general, our method achieves better accuracy and computational efficiency compared to standard and machine learning transfer learning methods on various tasks. Upon testing our method for populations with different insurance plans, we detect various levels of disparities in hospital duration stay between groups. By leveraging tools from OT theory, our proposed framework allows to compare statistical models on EHR data between different population groups. As a potential application for clinical decision making, we quantify treatment disparities between different population groups. Future directions include applying *OTTEHR* to broader regression and classification tasks and extending the method to semi-supervised learning.

**Data and Code Availability:** This paper uses the MIMIC-III dataset [Johnson et al., 2016], which is available on the PhysioNet repository [Moody et al., 2001]. The anonymized code repository is available at this link.

**Institutional Review Board (IRB):** This research does not require IRB approval.

## Introduction

An Electronic Health Record (EHR) database is a digital platform that securely stores and manages comprehensive health information for individual patients, offering healthcare providers quick and efficient access to crucial medical data. Building a comprehensive and unbiased database of EHRs is a crucial first step to precision and personalized medicine [Allen et al., 2012]. Comprehensive EHR databases generally achieve higher accuracy, avoiding potential issues of duplicate records and providing transparency among healthcare professionals, even across different healthcare databases [Menachemi and Collum, 2011]. Not just providing accurate information for each patient, a compendium of EHRs can serve as an important data set for augmenting human intelligence so that medical professionals can make informed decisions in everyday practice [Wagner et al., 2020].

In practice, it has been suggested that each EHR database was built for a different purpose to maximize its utility based on the needs [Kashani and Herasevich, 2015], making certain medical conditions highly, if not only, prevalent in specific studies and population groups [Woolf et al., 1955]. Having such an unequal distribution of medical conditions across different databases impedes our ability to diagnose rare conditions isolated within a specific population group. For example, alpha-1 antitrypsin deficiency, which affects the lungs and liver, is relatively uncommon in the general population but has higher prevalence rates in certain ethnic groups, such as those of Northern European descent [de Serres et al., 2007]. The variation in prevalence poses diagnostic challenges for patients in areas where the condition is less common. Moreover, unlike any other field of data science, data integration is practically not an option due to patient privacy and some hidden interests of stakeholders [Haas et al., 2011], which makes this problem more challenging.

Overcoming this challenge introduces the need to transfer knowledge learned from data-rich population groups to data-rare population groups, which can be expressed as unsupervised *transfer learning* (TL) [Ganin and Lempitsky, 2015], in the case where labels are only available for the data-rich population group. In this paper, we introduce ***O****ptimal* ***T****ransport-based* ***T****ransfer learning for* ***E****lectronic* ***H****ealth* ***R****ecords, OTTEHR*, a novel method that leverages feature embeddings from EHRs and computational Optimal Transport (OT) to perform unsupervised TL between unbalanced domains. As demonstrated in our application, this method allows us to compare the outputs of a model for different population groups, and in particular, quantify treatment disparities that can provide insightful guidance for clinical decision-making. To summarize, our contributions are as follows:

1. We introduce *OTTEHR* to enable unsupervised TL by applying barycentric projection from OT between unbalanced domains. Using experimental data from the MIMIC-III, MIMIC-IV and eICU databases, we show that in general, our method outperforms standard and recent unsupervised TL methods with respect to accuracy and computational efficiency.
2. We establish a theoretical upper bound for the generalization error of *OTTEHR* and decompose it into a source error and labeling divergence terms that are universal, and a specific transport term, that allows us in practice to assess the suitability of our method on specific datasets.
3. Upon applying *OTTEHR* in the context of predicting patient duration in hospital using medical codes across different groups (e.g insurance), we show that our method allows us to quantify treatment disparities, suggesting potential applications to uncover treatment biases among subgroups and improve patient care.

## Background and Significance

### Background

In this section, we introduce elements of OT and TL used in our study.

### Optimal Transport

OT aims to solve a general transport problem where we consider moving data points in one distribution of mass to another at a minimal cost. In its discrete version, the OT problem can be formulated as follows: Consider two distributions *µ*_*A*_ and *µ*_*B*_ of point support **A** = {**a**_*i*_ ∈ ℝ^*k*^, *i* = 1, …, *n}* and **B** = {**b**_*i*_ ∈ ℝ^*k*^, *i* = 1, …, *m*} with probability mass functions *ϕ*_*A*_ and *ϕ*_*B*_, respectively. We define a cost function *d* : **A***×* **B**→ ℝ_≥0_ (e.g. the Euclidean distance between **a**_*i*_ with **b**_*j*_), and the associated objective function

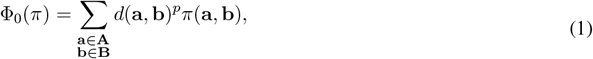

where *π* ∈ Mat_*m*,*n*_(ℝ_≥0_) (the set of *m × n* matrices with entries in ℝ_≥0_) and *p* ∈ [1, +∞]. Minimizing this objective function with the marginal constraints 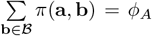 and 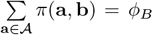 defines the classical OT problem.

To deal with the computational cost of the classical OT formulation and handle distributions with different mass, *unbalanced entropy-regularized OT* was recently introduced [Pham et al., 2020] by adding additional regularization constraints to Equation 1 as

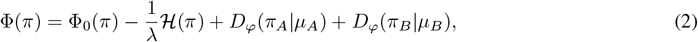

where 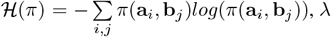 is the regularization parameter, *π*_*A*_ = *π***1**_*n*_ (**1**_*n*_ is the identify matrix of *n×n*), and *π*_*B*_ = *π*^⊤^**1**_*m*_; *D*_*φ*_(*α, β*) is the Csiszár *φ*−divergence and is given by, assuming that the discrete measures 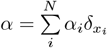 and 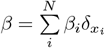 share the same support 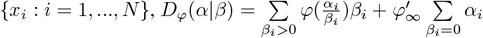 where 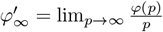.

Minimizing φ leads to define the *regularized unbalanced p-Wasserstein distance*, as

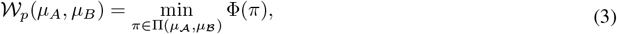

where Π is the set of positive matrices (that in this context define transport plans between *µ*_*A*_ and *µ*_*B*_), 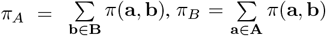, and the minimizer of Equation (3) gives the *OT plan*. In the rest of the manuscript, we will work with *p* = 1, and will simply refer to the regularized unbalanced 1-Wasserstein distance as the *Wasserstein distance* for simplicity.

Upon finding *π*\*, and assuming 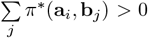 (so some mass gets transported from **a**_*i*_), the transported features of **a**_*i*_ can then be derived by *barycentric projection*, as

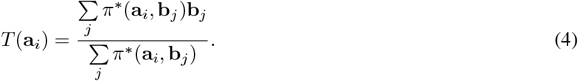

*Gromov-Wasserstein OT* extends the OT framework to the case where the cost function depends on the metrics associated with each distribution. The objective function for *regularized unbalanced Gromov-Wasserstein OT (UGWOT)* is:

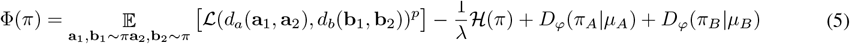

where ℒ: ℝ_≥0_ *×* ℝ_≥0_ → ℝ_≥0_ is a cost function on ℝ_≥0_, and *d*_*A*_ : **A** *×***A** → ℝ_≥0_ and *d*_*B*_ : **B** *×* **B** → ℝ_≥0_ are cost functions on **A** and **B**, respectively.

### Theoretical Framework for Transfer Learning

We extend the theoretical framework of TL for binary classification [Ben-David et al., 2010] to include regression tasks as follows: Consider two spaces, a *source space*, represented by a distribution *µ*_*S*_ on *source domain D*_*S*_ and a labeling function *f*_*S*_, and a *target space*, represented by a distribution *µ*_*T*_ on *target domain D*_*T*_ and a labeling function *f*_*T*_. The label domain is {0, 1} for binary classification and ℝ for regression. We define a binary classifier/regressor *h* : *D*_*S*_ →{0, 1} or ℝ, so the probability that the binary classifier/regressor *h* disagrees with a labeling function *f* according to the distribution *µ*_*S*_ is 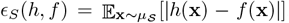,where |·| is the *l*_1_ norm metric on ℝ. In particular, the source error of *h* is *ϵ*_*S*_(*h*) = *ϵ*_*S*_(*h, f*_*S*_); and similarly, we define the target error, also known as the generalization error of *h*, as

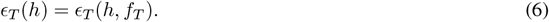

Then, the problem of TL via OT aims to fine-tune a regressor *h*\*, learned from the source domain, into *h*^*′*^ so that the resulting target error *ϵ*_*T*_ (*h*^*′*^) is minimized.

### Significance

Our study provides an OT-based TL framework for EHR data with potential applications on detecting treatment disparities, as shown in the interdisciplinary pipeline in Figure 1.A. The framework of the method can be found in Figure 1.B. Below are some more details on the novelty of our study regarding the application of OT-based TL and theoretical guarantees.

**Figure 1:**
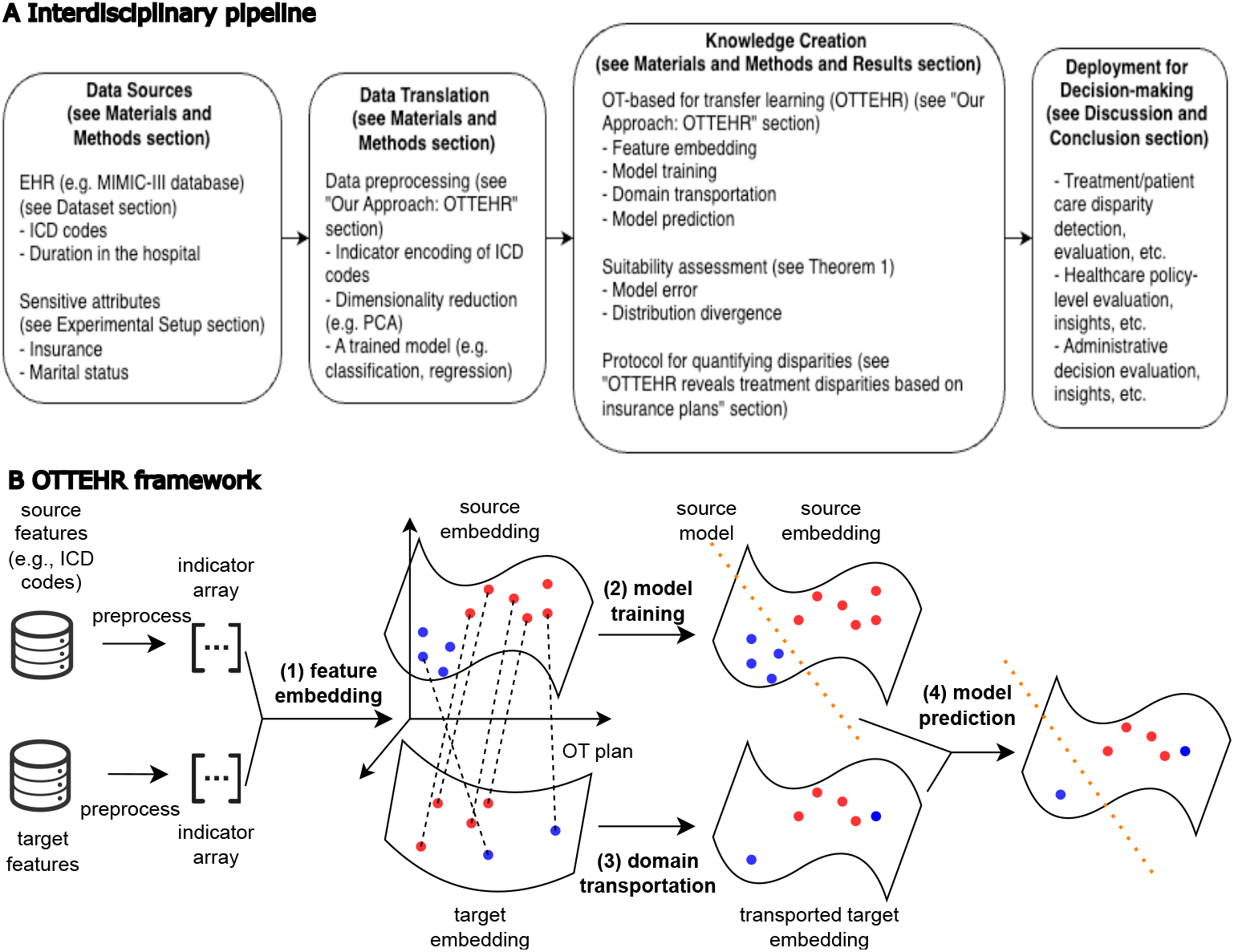
Interdisciplinary pipeline and framework for *OTTEHR*. (**A**) Interdisciplinary pipeline for *OTTEHR*. (**B**) The framework of *OTTEHR*. The four major steps, (1) feature embedding, (2) model training, (3) domain transportation, and (4) model prediction, are in bold. The orange dotted lines denote the trained source classifier/regressor from step (2). The black dotted lines denote the OT plan learned from step (3). The colors of the points on the embedding spaces represent different classes. Note that when using *OTTEHR* the labels for the target features are not required and they present here for the visualization purpose.

### Application of OT-based Transfer Learning in Healthcare

In healthcare, OT-based TL has been applied to sepsis prediction from EHRs [Ding et al., 2023, Wang et al., 2022], mapping prostate cancer across MRI scanners [Gautheron, 2017], enhancing EEG-based motor imagery recognition [Chen et al., 2022], and improving P300 detection for brain-computer interfaces [Liu et al., 2020]. However, other variables, such as medical codes, can also introduce distributional shifts, posing challenges for transfer learning. Our study studies some examples of transfer learning tasks that involve medical codes with distributional shifts, either among subgroups within single or among multiple healthcare databases. In addition, our framework can be further utilized for downstream applications such as the quantification of treatment disparities. In summary, our proposed framework is designed to *fl*exibly facilitate and analyze transfer learning tasks involving various types of variables across healthcare databases.

### Error Upper Bound for OT-based TL

Ben-David *et al*. defined a formal model of TL, also known as domain adaptation, in the case of binary classification [Ben-David et al., 2006], and derived a theoretical upper bound on the error of target data [Ben-David and Urner, 2014, Ben-David et al., 2010]. We extend the formal model of TL to regression tasks and prove the upper bound on the error of the target data in the context of unbalanced OT. Courty *et al*. extended these concepts to explore the target error bound of balanced OT in a joint training process [Courty et al., 2017]. In contrast, our work focuses on unbalanced OT, with a two-phase training process for the model training and domain transportation. Separating the training into two phases can allow each loss function to be optimized more efficiently [Malik et al., 2021, Li et al., 2022], while using “unbalanced” OT can improve the robustness of our framework in the context of EHRs, where datasets of different groups are rarely balanced.

## Materials and Methods

### Dataset

The MIMIC databases, including MIMIC-III and MIMIC-IV, provides comprehensive, open-source, de-identified health-related data for patients admitted to the Beth Israel Deaconess Medical Center [Johnson et al., 2016, 2020]. The MIMIC-III database contains over 40,000 patients and 58,976 admissions between 2001 and 2012. It is a relational database consisting of 26 tables consisting of 6,918 unique ICD-9 diagnosis codes. The MIMIC-IV database contains over 65,000 patients admitted to an ICU and over 200,000 patients admitted to the emergency department with 76,540 admissions, consisting of 29 tables with 7,942 unique ICD-10 diagnosis codes. We focused on the patient tables, admission and diagnosis tables. In the patient table, each patient is attached with patient ID, date of birth and insurance. In the admission table, each admission is attached with the admission ID, patient ID, admission time, and discharge time. In the diagnosis table, each diagnosis is attached with the admission ID and the diagnosed ICD code.

The eICU Collaborative Research Database is a multi-center database comprising de-identified health-related data associated with over 200,000 admissions to intensive care units (ICUs) across the United States collected between 2014 and 2015 Pollard et al. [2018]. The eICU database contains data from more than 139,000 patients from 335 hospitals, consisting of 31 tables with 857 unique ICD-9 diagnosis codes. We focused on the patient and diagnosis tables. In the patient table, each patient is attached with admission ID, hospital ID, admission and discharge time. In the diagnosis table, each diagnosis is attached with the admission ID and the diagnosed ICD codes.

### Our Approach: *OTTEHR*

We propose our method *OTTEHR*, short for *Optimal Transport-based Transfer Learning for Electronic Health Records*, to address TL problems in EHR data using OT. We set up our notations as follows. Let χ be the input domain and 𝒴 be the label domain. Let (**X**^*S*^, **Y**^*S*^) denote the pair of source features and source labels and (**X**^*T*^, **Y**^*T*^) denote the pair of target features and target labels, such that 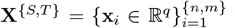 where *n* and *m* are the numbers of points in source and target domains and {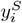 ∈ ℝ or ∈ {0, 1}}. We preprocess the numerical medical codes into indicator arrays so that the *l*^*th*^ coordinate in **x**_*i*_ denotes the presence of the *l*^*th*^ code, using 0 or 1. We assume the unknown target labels **Y**^*T*^ are also in the same space as **Y**^*S*^.

*OTTHER* proceed in four main steps, also shown in Figure 1.B:

#### (1) Feature embedding

Feature embedding is important to process EHRs as *(i)* EHRs are of high dimension, and the classifier/regressor *h*\* in step (2) and the OT plan *π*\* in step (3) cannot be learned well without proper dimensions; *(ii)* EHRs are also sparse, and feature embedding can be done effectively to resolve *(i)*.

We run Principle Component Analysis (PCA) Jolliffe [1990] to embed important information into low-dimensional vectors on source features, that yields a mapping 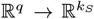 where *k*_*S*_≪ *q*, where *k*_*S*_ is chosen to explain approximately 75% of variances in source domains. Applying *g*_*S*_ to source features yields source embedding as

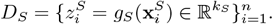

For target embeddings, two scenarios are considered:

First, when source and target features share the same space (e.g., ICD codes from the same hospital database), applying *g*_*T*_ = *g*_*S*_ to target features yields target embedding as

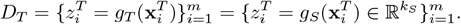

Second, when source and target features do not share the same space (e.g., different versions of ICD codes from different hospital databases), we run a separate PCA to embed important information into low-dimensional vectors on target features, producing a mapping 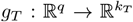 where *k*_*T*_ ≪ *q*, where *k*_*T*_ is chosen to explain approximately 75% of variances in target domains. Applying *g*_*T*_ to target features yields target embedding as

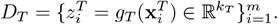

Note that our procedure allows to replace PCA by more advanced dimensionality reduction methods, as mentioned in the Discussion section.

#### (2) Model training

We train a classification or regression model *h*\* : *D*_*S*_ → *Y*_*S*_ using the source embedding *D*_*S*_ and source labels *Y*_*S*_.

#### (3) Domain transportation

In parallel of the model training step, we learn an OT plan *π* between the source and target as defined in Equations (2) and (3) or Equations (3) and (5), and use it to map all target embeddings in *D*_*T*_ onto the source embedding domain *D*_*S*_ using equation 4, so the set of transported target embeddings is

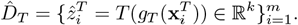

#### (4) Model prediction

The source model is applied to the transported target embedding to obtain the “projected” target labels. We predict the label, 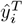 for all 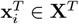,using

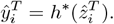

### Implementation Details

We implemented *OTTEHR* in Python 3, with the source code available at this link. To solve OT problems, our package depends on the *POT* library [Reinger, 2021]. We ran our experiments on a workstation with 32 central processing units powered by AMD Ryzen Threadripper 2950X 16-Core Processor, 125GB of RAM, and x86_64 Ubuntu 20.04.5 LTS. We used Euclidean distance (*l*_2_ norm) as the cost function for OT and Kullback-Leibler divergence as the mass divergence *D*_*φ*_. For OT hyperparameters, we set the entropic regularization parameter *λ* to 0.1 and the mass regularization parameter to 1. These hyperparameters can be selected using cross-validation by splitting a small set of data into training and validation sets, then optimizing these parameters to minimize a predefined validation loss.

### Experimental Setup

In the following, we describe the common data processing and experimental procedures across all four experiments, and detail the differences in group division and additional analysis for each experiment.

For all experiments, we ran *OTTEHR* using linear regression models to predict duration in hospitals. For each pair of source and target groups, we randomly sampled 120 admissions (training) from the source group and 100 (testing) admissions from the target group, and we conducted the same type of experiment repeatedly 100 times. The number of ICD codes varies from 700 to 900 for each experiment, depending on the selected admissions. During the feature embedding step, we convert the explanatory variables into a 50-dimensional space (*k* = 50). We benchmarked *OTTEHR* against existing TL methods using *mean absolute error (MAE)* and *root mean squared error (RMSE)*. For details on data preprocessing for each experiment and evaluation metrics, please see Section 1 experimental details section in SI.

### Insurance experiments

We divided the MIMIC-III admissions into source and target by insurance groups. Insurance groups include “Self_Pay,” “Private,” “Government,” “Medicare,” “Medicaid.” We ran *OTTEHR* on all possible pairs, considering one group as source and the other as target, resulting in 20 (i.e., 5 *×*4, where 5*×* 4 represents the number of source-target pairs within the five insurance groups) distinct experiments. We treated the presence of each ICD code as the explanatory variable. We confirmed the existence of distributional shifts in explanatory variables between pairwise insurance groups by visualizing the PCA embeddings of explanatory variables in Figure S1 in SI, showing the differences in the feature embedding space between each pair of source and target groups are significant.

In Table S1 of SI, we further confirmed the distributional shifts between certain group pairs by Mann-Whitney U tests McKnight and Najab [2010], with the p-values smaller than 0.05 indicating significant differences. In addition to benchmarking against existing TL methods, we analyzed the empirical relationship between the target error of *OTTEHR* and other terms presented in the upper bound of the target error in the Results section, and applied *OTTEHR* to detect treatment disparities across different insurance plans.

### Cross-database experiments

We considered the MIMIC-IV (which uses ICD-10) as the source group and MIMIC-III (which uses ICD-9) as the target group. We treated the presence of each ICD code as the explanatory variable. Like in the insurance experiments, We confirmed the distributional shift by visualizing the PCA embeddingsin Figure S2 in SI, and by finding significant p-values from Mann-Whitney U tests in Table S2 in SI.

### Age experiments

We divided the MIMIC-III admissions into the following age groups [25, 45], [30, 50], [35, 55], [45, 65], [50, 70], [55, 75], and [60, 80]. We considered [50, 70] as the source and other groups as the target groups, which results in six experiments. We treated the presence of each ICD code and age as the explanatory variables. Since it is clear that the distributional shifts in this example are introduced by continuous age feature, there is no need to visually confirm the distributional shifts as in the previous experiments.

### Cross-hospital experiments

We divided eICU admissions into source and target by hospital IDs. We chose the top 10 hospitals with the most admissions. These hospital IDs are 420, 264, 243, 338, 73, 458, 167, 443, 208 and 300. We ran *OTTEHR* on all possible pairs, resulting in 90 distinct experiments. We treated the presence of each ICD code as the explanatory variable. The distribution shifts between different hospitals have been confirmed by existing literature Rajabalizadeh et al. [2020]. Like for the insurance and cross-database experiments, we confirmed the distributional shifts by visualizing the PCA embeddings in Figure S3 in SI and calculating p-values from Mann-Whitney U tests in Table S3 in SI, and noted that they are more pronounced than in the insurance and cross-database experiments.

## Results

### Upper Bound on the Target Error for Binary Classification and Regression

To study the theoretical accuracy of *OTTEHR*, we derived an upper bound on the target error in the case of binary classification and regression. Our analysis focuses on Lipschitz continuous models. In practice, many models, including for EHRs, satisfy this condition [Goldstein et al., 2017, Clegg et al., 2016, Harutyunyan et al., 2019] or can be approximated as Lipschitz continuous [Bartlett et al., 2017, Nair and Hinton, 2010, Ioffe and Szegedy, 2015], and assuming Lipschitz continuity or linearity is common in theoretical studies of TL [Tong et al., 2021, Tian and Feng, 2023, Cai et al., 2021].

As we adapt embeddings from the target domain to the source domain using the barycentric operator *T*, the fine-tuning of the learned function on the source *h*\* yields *h*^*′*^ = *h*\* *T*. Assuming 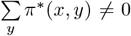 (see Equation (4)), we can control the target error by the following theorem.

#### Theorem 1

(Upper bound for target error). *Let µ*_*T*_ *be a discrete target distribution defined on a domain D*_*T*_ *and with probability mass function ϕ*_*T*_, *and µ*_*S*_ *be a discrete source distribution defined on a domain D*_*S*_ *and with probability mass function ϕ*_*S*_. *Let h*^*′*^ = *h*\* ○ *T*, *where h*\* *is Lipschitz continuous and T is the barycentric projection (Equation* (4)*). The target error ϵ*_*T*_ (*h*^*′*^) *defined by Equation* (6) *is bounded by:*

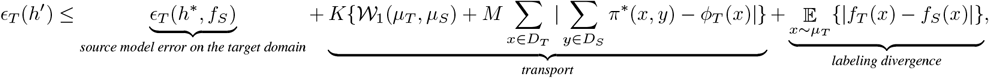

*where K is the Lipschitz continuous constant for h*\*, *π*\* *is the OT plan and* 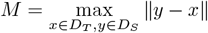

The proof can be found in Section 2 of SI. We interpret the upper bound as follows. The first term is the source model error evaluated on the target domain, illustrated by Figure S4 in SI. The second “transport term” is composed of two p arts: *(i)* 𝒲_1_(*µ*_*T*_, *µ*_*S*_) that denotes the Wasserstein distance between *µ*_*T*_ and *µ*_*S*_ (see Equation (3)), and *(ii)* 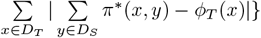 that denotes the variance between mass at *x* and the transported mass from *x* to *D*_*S*_, indicating the “unbalancedness” of the OT plan (the larger *D*_*ϕ*_(*π*_*A*_|*µ*_*A*_) and *D*_*ϕ*_(*π*_*B*_|*µ*_*B*_) in Equation (2), the larger this term will be). The last “labeling divergence” term 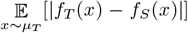 denotes the divergence between the source and target true labeling functions.

Note that when OT is used rather than unbalanced OT, we have the marginal constraints 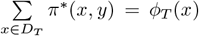 resulting in eliminating unbalancedness term from Theorem 1. Also note that when *h*\* is linear, a special case of Lipschitz continuity, *K* in the theorem can be substituted by 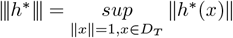.

This generalization error bound provides a guide for analyzing and interpreting the performance of our method. More precisely, if the transport term is significantly smaller compared to the source model error and labeling divergence, then using OT to map individuals to another group should be a suitable option for the TL task. Note that the estimation of the labeling divergence requires access to ground-truth target labels 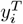 to estimate the labeling function on the target domain *f*_*T*_, which might not be available in practice. Nevertheless, we can mitigate this issue with partial access to the target labels or prior knowledge of *f*_*T*_.

### Application of the Upper Bound

Having established the theoretical framework and derived an upper bound for the target error in the previous section, we now explore its practical implications and validate its utility using insurance experiments. More precisely, we focused on how the duration of hospitalization can be predicted by medical codes for different insurance groups (as detailed in the Materials and Methods section), as a simple but relevant test case [Goldman and Smith, 2002, Umberson, 1987]. We compared the target errors of *OTTEHR* versus other terms presented in Theorem 1. Note that in this context, using the same model for estimating 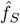 as *h*\* leads to 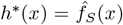 for all *x* ∈ *D*_*T*_, and 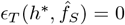,so it was not necessary to study the relationship between the target error and the source model error on the target domain, leaving the transport term and the labeling divergence to be studied.

We separately plotted in Figure 2 the target error against the transport term and against the labeling divergence with the results for all the insurance group experiments combined. We observed a strong correlation between the target error and labeling divergence, with a Pearson correlation coefficient (PCC) of 0.70 (Figure 2.A), in contrast to a weaker correlation between the target error and the transport term, with a PCC of 0.16 (Figure 2.B). We explain these results by the large difference in orders of magnitude between the labeling divergence (10^8^) and the transport term (10^0^). Further analysis of pairwise insurance groups confirmed consistent trends (see Figure S5 and S6 in SI), indicating that in our experiments, the target error is overall dominated by the labeling divergence. As previously discussed in the Results section, since the transport term is significantly smaller than the labeling divergence, *OTTEHR* is suitable for solving these TL tasks.

**Figure 2:**
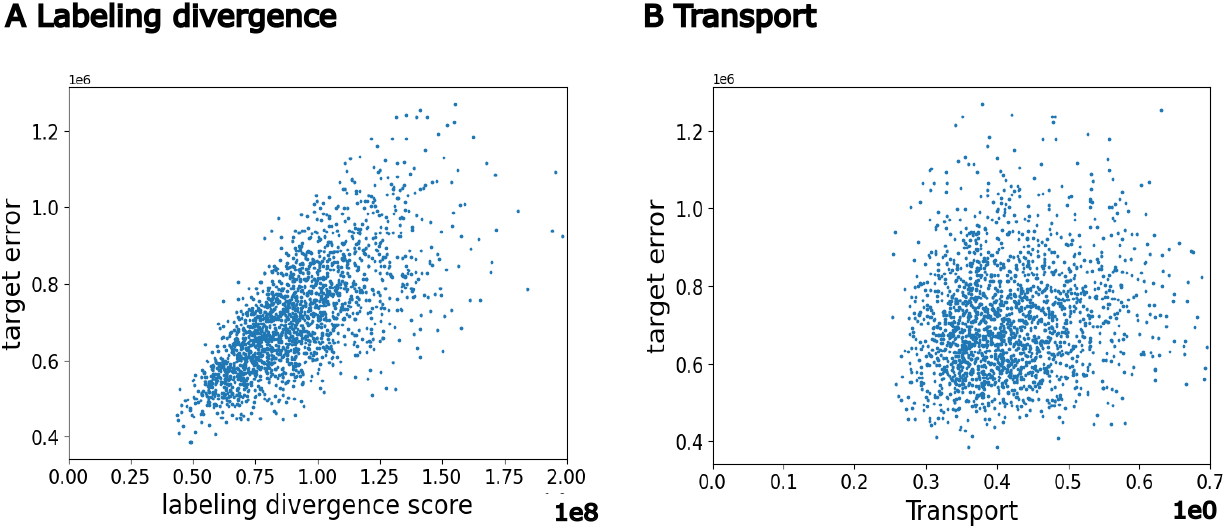
Bound analysis for all insurance group experiments. The relationship between target error and (**A**) labeling divergence term with a PCC of 0.70, and (**B**) transport term with a PCC of 0.16, combining the results for all pairwise insurance group experiments.

### Benchmarking of Accuracy and Computation Time

To comprehensively evaluate the performance of *OTTEHR*, we assessed its accuracy and computation time for all three experiments.This evaluation provides insights into *OTTEHR*’s effectiveness under diverse scenarios, including distributional shifts caused by continuous (i.e., age) or discrete features (i.e., insurance, ICD version), varying degrees of distributional divergence (i.e. age experiments), and transfer learning tasks conducted within a single database (i.e., insurance experiments) or across multiple databases (i.e., cross-database experiments).

In Section 3 of SI, we provide a brief overview of standard statistical and machine learning based methods used for TL. To assess the empirical performance of *OTTEHR*, we then opted to benchmark it against Transfer Component Analysis (*TCA*) [Pan et al., 2010], Correlation Analysis (*CA*) [Sun et al., 2017], Geodesic Flow Kernel (*GFK*) [Gong et al., 2012], Deep Joint Distribution Optimal Transport (*deepJDOT*) [Damodaran et al., 2018], Representation Subspace Distance (*RSD*) [Chen et al., 2021] and inverse GRAM matrices (*daregram*) [Nejjar et al., 2023], as these methods provide a good representation of the different methods described in SI. We benchmarked the aforementioned methods for all experiments (for more details on the protocol and metrics used (mean absolute error (*MAE*) and root mean square error (*RMSE*) [Chai and Draxler, 2014]), see Material and Methods).

For insurance, cross-database and cross-hospital experiments, violin plots in Figure 3.A to F shows the log-transformed *MAE* and *RMSE*, with smaller values indicating better performance. We notably observed that *OTTEHR*’s median *MAE*/*RMSE* is smaller than those of *TCA, CA, GFK, RSD* and *daregram* with comparable standard deviations. Although *OTTEHR*’s median *MAE*/*RMSE* is slightly larger than that of *deepJDOT* for insurance and cross-database experiments, its standard deviation is significantly smaller. We also provided detailed results in Table S4-S11 in SI for pairwise insurance, MIMIC-IV and MIMIC-III, and pairwise hospital groups, showing the medians and standard deviations of *MAE*/*RMSE* for all the methods, and outperformance ratios of *OTTEHR* to other methods. Overall, *OTTEHR* achieved a 8.61% to 53.73% reduction in median *MAE/RMSE* compared to *CA, GFK* and *RSD* and *daregram* and a 36.67% to 89.87% reduction in the standard deviation of the log-transformed *MAE/RMSE* compared to *deepJDOT*. In addition, *OTTEHR* achieved a comparable performance to *TCA* on cross-database and cross-hospital experiments, and a 25.17%/14.79% reduction in median *MAE/RMSE* compared to *TCA* in insurance experiments.

**Figure 3:**
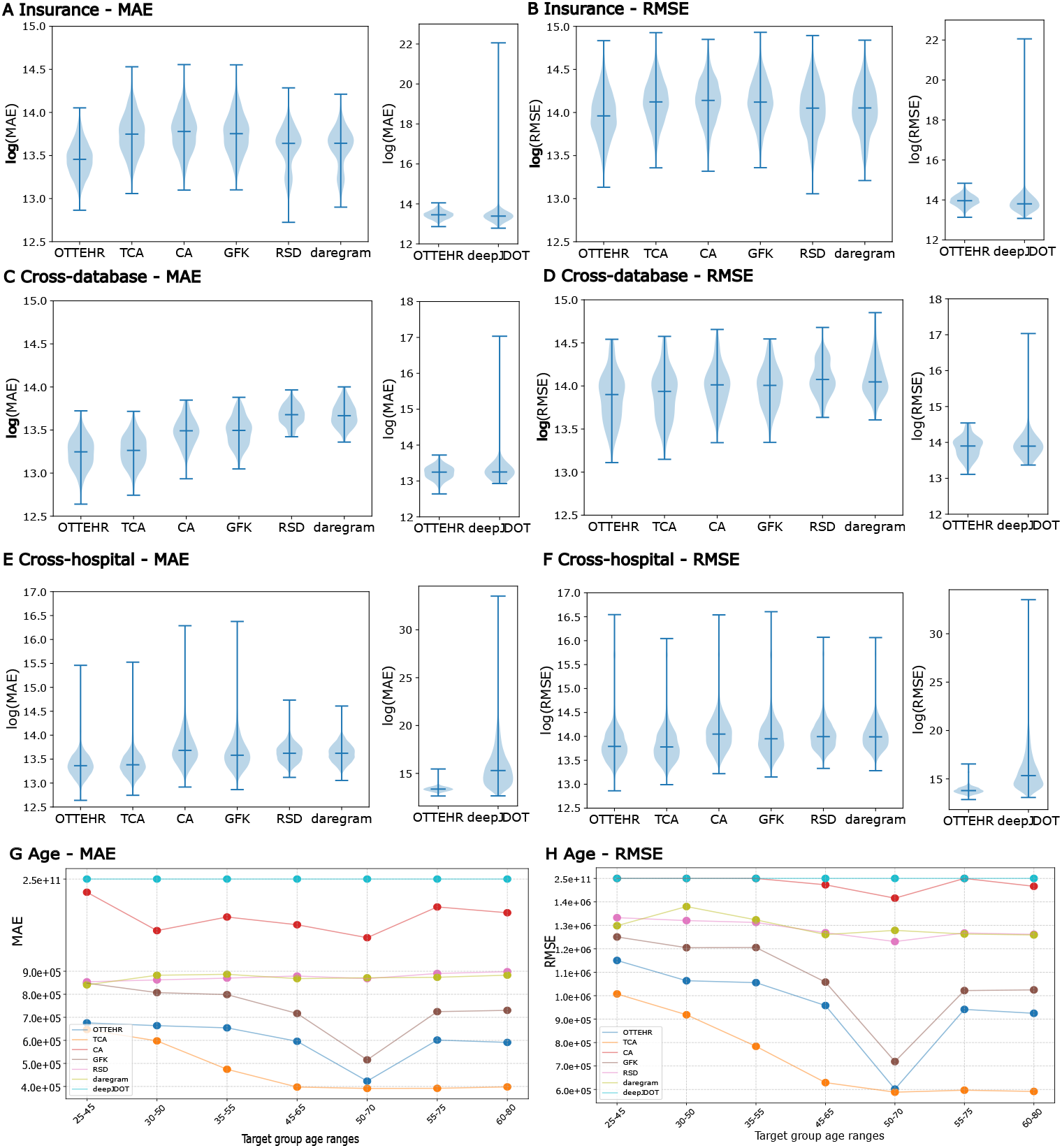
Benchmark results of *OTTEHR* for all experiments against competing methods. Violin plots of the log of (**A**)/(**C**)/(**E**) *MAE* and (**B**)/(**D**)/(**F**) *RMSE* between projected duration and observed duration on target admissions for insurance/cross-database/cross-hospital experiments. The blue bars denote the medians. The heights of the violin plots denote the variability, positively correlated with the standard deviations. Smaller *MAE* and *RMSE* values indicate better performance. Scatter plots of median (**G**) *MAE* and (**H**) *RMSE* between projected duration and observed duration on target admissions for age experiments.

For the age experiments, Figure 3.E and F show median *MSE* and *RMSE* across target groups in different age ranges. We observe that as the overlap in age between the source and target groups increases, the performance of *OTTEHR* improves, achieving its best *MAE*/*RMSE* when the source and target groups fully overlap in the age range [50, 70]. Moreover, *OTTEHR* outperforms methods such as *CA, GFK, RSD, daregram*, and *deepJDOT*, showcasing greater robustness by relying less on substantial distributional overlap for strong performance. While *TCA* consistently outperforms *OTTEHR* across all age experiments, this can be explained by *TCA*’s ability to identify a shared meaningful space between source and target groups when distributional shifts are minimal particularly those induced by continuous features such as age in the age experiments. Notably, as distributional shifts become more significant (e.g., when the target group ages fall within [20, 40]), the performance of *TCA* and *OTTEHR* converges, indicating *OTTEHR*’s robustness under significant distributional shifts.

In addition to accuracy, we compared *OTTEHR* with competing methods in terms of average computation time per experiment. In Table S12 in SI, we observe in average, *OTTEHR*’s runtime performs better (on the same order of magnitude) than *TCA, GFK, RSD* and *daregram*, 28 (9786*/*348) times faster than that of *CA*, and 48 (16603*/*348) times faster than that of *deepJDOT*.

To conclude, *OTTEHR* overall consistently demonstrates superior performance in accuracy and in computational efficiency compared to its competitors across all four experiments.

### Application to Detect Treatment Disparities

After confirming the empirical performance of *OTTEHR*, we applied it to quantify treatment disparities based on insurance plans and predict the potential impact of transitioning between different plans. We focused on Medicaid, a collaborative program that assists individuals with limited income and resources [Gruber, 2003], Medicare, that provides coverage to people aged 65 and older, as well as to younger individuals with certain disabilities and diseases [Finkelstein and McKnight, 2008], and Private insurance, offered by various companies, that offers greater *fl*exibility in healthcare services [Cutler and Gruber, 1997]. People often switch from Medicaid and Medicare to private insurance due to factors such as reductions in federal matching funds or when their preferred healthcare providers fall out of their Medicare network [Foundation, 2023, Kaiser Family Foundation, 2023, 2021]. Conversely, transitions to Medicaid and Medicare from private insurance can result from changes in age, income, or health status Long et al. [2014], Baicker and Chandra [2006].

Specifically, we considered the projected duration obtained from TL to be significantly reduced if it is at least 300 hours (12.5 days) shorter than the originally observed duration. Note that we focused on significantly reduced durations rather than all reduced durations, ensuring that the findings we report are statistically robust and not due to random variation. In Figure 4, we showed kernel density estimate plot (KDE) of projected duration versus the original duration in hours for admissions transitioning (**A**) from Medicaid to private insurance, (**B**) from private insurance to Medicaid, (**C**) from Medicare to private insurance, and (**D**) from private insurance to Medicare, with blue dots denoting admissions with significantly reduced durations, where we observed very different proportions of admissions with significantly reduced durations. We note that the significantly reduced admissions in transitions from Medicaid/Medicare to private and from private to Medicaid/Medicare are distinct subsets of the population. Hence, the presence of significantly reduced admission in one direction does not indicate that there is no significantly reduced admission in the other direction. Our findings indicate that 13.1% of admissions would result in significantly reduced durations when transitioning from Medicaid to private insurance, compared to only 9.5% with such reductions when transitioning from private insurance to Medicaid, making a difference of 3.6%, where the difference adjusts for distributional shifts between different pairwise insurance groups. Similarly, 9.0% of admissions would result in significantly reduced durations when transitioning from Medicare to private insurance, compared to 10.5% with such reductions when transitioning from private insurance to Medicaid, making a difference of 1.5%. The larger difference of bi-directional transition process between private insurance and Medicaid suggests more disparities between private insurance and Medicaid compared to those between private insurance and Medicare. Such results can be generalized to more insurance groups (e.g., Government), as shown in Figure S7 in SI, where percentages of admissions with significantly reduced durations are shown across all pairs of insurance groups.

**Figure 4:**
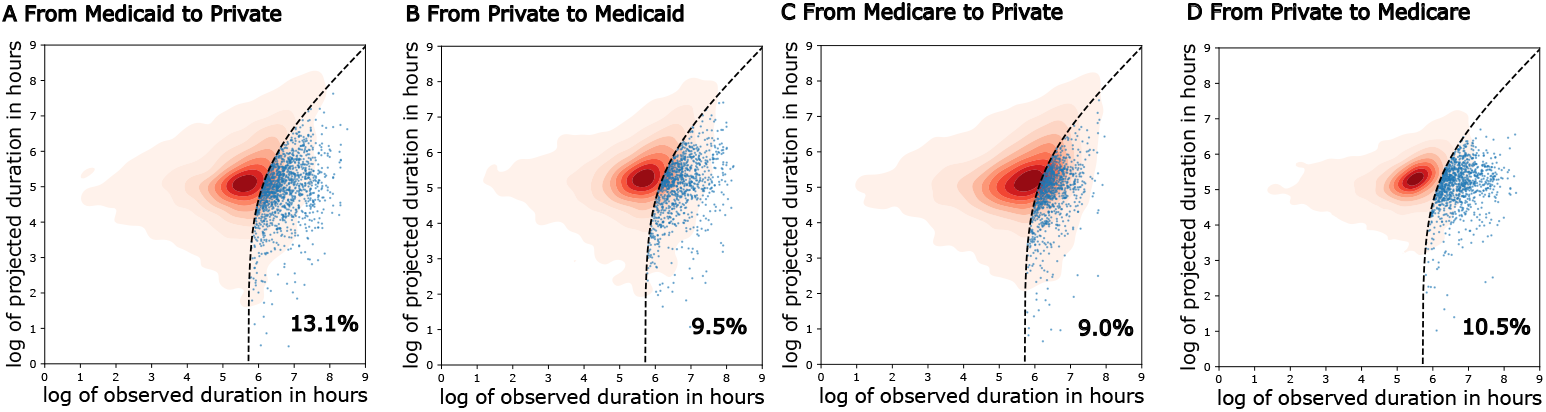
Admissions with significantly reduced durations of stay in hospital when transitioning between private insurance and Medicaid/Medicare. The KDE plot of the log of projected duration in hours versus the log of observed duration in hours for admissions transitioning (**A**) from Medicaid to private insurance, (**B**) from private insurance to Medicaid, (**C**) from Medicare to private insurance, and (**D**) from private insurance to Medicare. The dotted black lines denote projected duration = observed duration − 300. The blue dots denote admissions with significantly reduced duration of stay in hospital, where the projected duration is at least 300 hours (12.5 days) less than the observed duration. The annotated percentages are the proportions of significantly reduced durations.

## Discussion and Conclusion

This paper presents *OTTEHR*, an OT-based unsupervised TL framework for EHRs. While biased models can lead to incorrect diagnoses, treatments, and healthcare decisions [Chen et al., 2023, Mittermaier et al., 2023], *OTTEHR* can potentially alleviate these biases by leveraging OT when comparing different population groups. Our study more precisely establishes a theoretical upper bound for the generalization error. Interestingly, we decomposed this bound into some general terms (namely the source error and the labeling divergence) that are shared by any TL method, and a specific transport term, that we can use in practice to evaluate the suitability of our method on real data, as shown in our application to the MIMIC-III dataset. We also note that all these terms are computable (as we did in Figure 2 and Figure S5-S6 in SI) or can be estimated if we have limited access to target labels or some prior knowledge about the target domain’s labeling function. Overall, our benchmarking suggests that in the context of EHRs, *OTTEHR* overall consistently demonstrates superior performance in accuracy and in computational efficiency compared to its competitors across all three experiments. Moreover, it is on par or performs better than *TCA* when the distributional shifts between source and target groups are sufficiently large. This observation is consistent with the fact that the Wasserstein metric in OT is a widely-used approach for addressing distribution shift problems in optimization Kuhn et al. [2019]. Upon focusing on predicting duration in hospital by transferring knowledge between insurance plans, we also detected significant differences underlying treatment disparities across insurance groups, suggesting our method’s potential for uncovering treatment biases among subgroups and helping in the clinical decision-making process for improved care.

To conclude, there are various potential future directions. It would first be interesting to further evaluate *OTTEHR* on other relevant regression and classification tasks and other demographic factors. These include predicting the time interval between consecutive visits [Poole et al., 2016] and mortality rates [Goodacre et al., 2006], using appropriate and potentially larger and local datasets [Pader et al., 2021, Sudlow et al., 2015]. As an effective and efficient TL method, *OTTEHR* could be applied to various challenges in clinical domain, such as generalizing early sepsis detection models Wang et al. [2022], Ding et al. [2023] and comparing surgical outcomes amongst surgeons from different subgroups, where the explanatory variables are not limited to medical codes Wallis et al. [2017]. More hypothetically, if paired with machine learning methods for phenotype clustering, such as those for COVID-19 pneumonia Lau et al. [2022], Gattinoni et al. [2020], *OTTEHR* can also rapidly compare and determine effective treatments. Furthermore, extending our method to semi-supervised TL [Wei et al., 2019] and designing a unified model that simultaneously solves feature embedding and classification problems [Song et al., 2017] could improve predictive performance on target domains with limited labeled data. From a theoretical perspective, it would be beneficial to extend the upper bound theorem, to include other tasks, such as, multi-class classification, non-continuous Lipschitz models, and other OT-metrics that can be more suitable for TL across different EHR databases [Séjourné et al., 2021]. Finally, there are several other potential areas for improving our current approach, including reducing its computational complexity to handle larger datasets, optimizing the embedding with more complex manifold learning techniques, and integrating heterogeneous information, such as laboratory results and doctor’s notes.

## Data Availability

All data produced are available online at https://github.com/wxli0/OTTEHR.

## Acknowledgments

We acknowledge the help from Zhenyuan Zhang and Greg d’Eon for discussing the proof for Theorem 1.

## Funding

This research received no specific grant from any funding agency in the public, commercial or not-for-profit sectors.

## Conflicts of Interest

The authors have no competing interests to declare.

## Author Contributions

Wanxin Li (Data curation, Formal analysis, Investigation, Methodology, Software, Validation, Visualization, Writing - original draft, Writing - review & editing), Saad Ahmed (Resources, Writing - review & editing), Yongjin P. Park (Conceptualization Formal analysis, Investigation, Methodology, Resources, Supervision, Validation, Visualization, Writing - original draft), Khanh Dao Duc (Conceptualization, Data curation, Formal analysis, Investigation, Methodology, Project administration, Resources, Supervision, Validation, Visualization, Writing - original draft, Writing - review & editing).

## Data Availability

The MIMIC datasets used in the experiments can be accessed upon request via PhysioNet. The source code for replicating the experiments is available at this GitHub repository.

## A. Experiment details

### A.1 Data preprocessing

For MIMIC-III/MIMIC-IV, we first merged the admission table with the patient table by indexing unique patient IDs. For each admission, we calculated the duration in the hospital by taking the difference between the discharge time and the admission time (in seconds). For the cross-database experiment, we append each admission with MIMIC version (i.e., MIMIC-III or MIMIC-IV). For age experiments, we calculated the age of the patient in each admission by taking the difference between the admission time and date of birth.

### A.2 Evaluation metrics

The evaluation metrics we used are *mean absolute error (MAE)* and *root mean squared error (RMSE). MAE* measures the average magnitude of errors between predicted and actual values, providing an intuitive sense of the typical prediction error in the same units as the target variable, which gives by

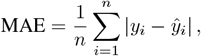

where *y*_*i*_ is the actual value, *ŷ*_*i*_ is the predicted value and *n* is the number of points. *RMSE* quantifies the standard deviation of prediction errors, emphasizing larger errors by squaring them, making it particularly sensitive to outliers.

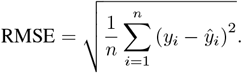

## B Proof of Theorem - Upper Bound for Binary Classification and Regression

Let *µ*_*T*_ (*µ*_*S*_) be a discrete target (source) distribution defined on a domain *D*_*T*_ (*D*_*S*_) and with probability mass function *ϕ*_*T*_ (*ϕ*_*S*_). Let *h*^*′*^ = *h*\* ○ *T*, where *h*\* is Lipschitz continuous and *T* is the barycentric projection (Equation 4 in the main manuscript). The target error *ϵ*_*T*_ (*h*^*′*^) defined by Equation 5 in the main manuscript is bounded by

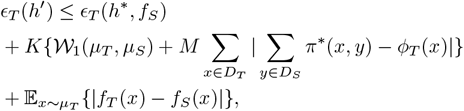

where *K* is the Lipschitz continuous constant for *h*\*, *π*\* is the OT plan and 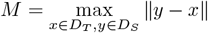.

*Proof*. We first rewrite the target error as

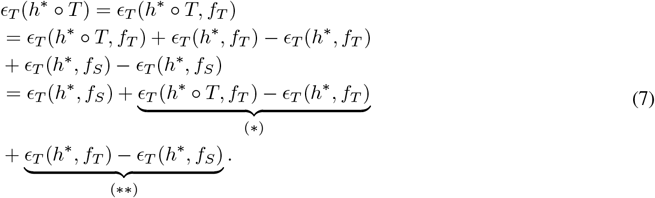

Since *K* is the Lipschitz constant for *h*\*, for all *x*, there exists *K >* 0 such that ∥*h*\* ○*T* (*x*) −*h*\*(*x*)∥ ≤ *K*∥*T* (*x*) −*x*∥. We now separately analyze (*) and (**). By the triangle inequality,

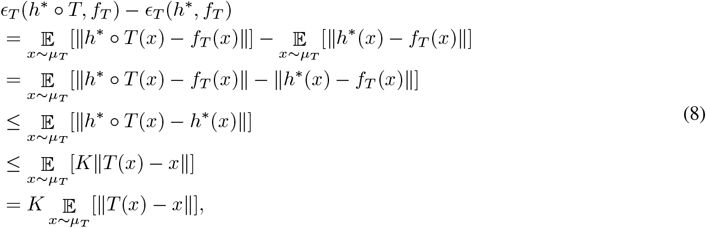

Let 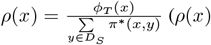 (is well defined since 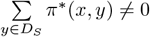)We then obtain

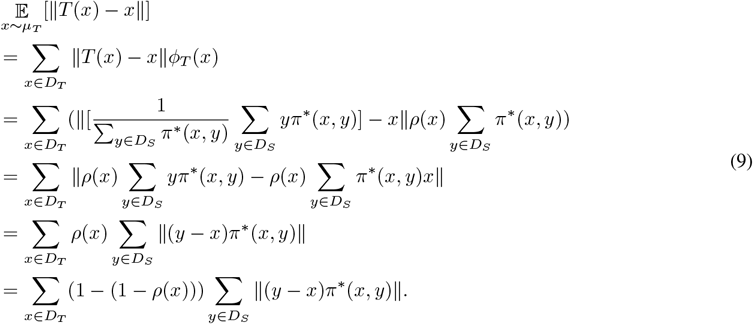

We can thus further bound 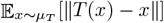 as

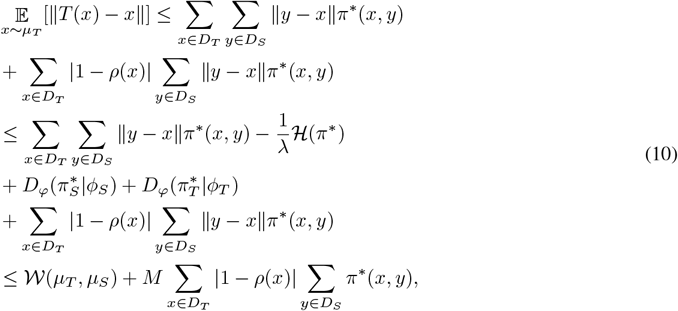

where 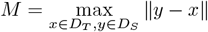. Finally,

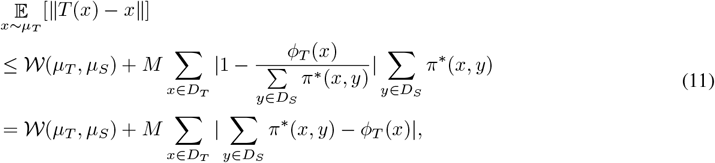

which yields the upper bound for (*)

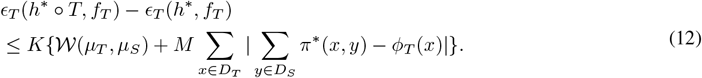

Considering (**), we have by triangle inequality,

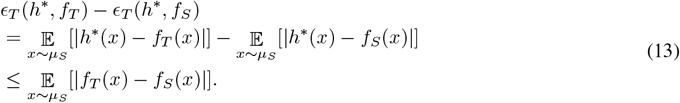

Plugging Equations (12) and (13) into Equation (7) yields

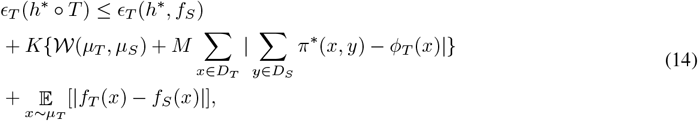

which completes the proof.

We note that when *h*\* is linear, Equation (8) can be rewritten in the following way:

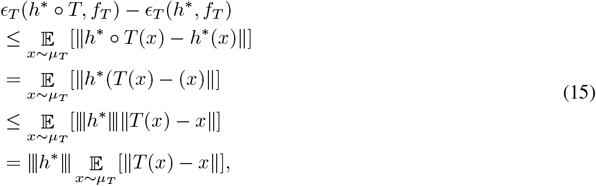

where 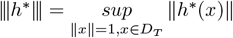.

In this case, the theorem can be rewritten as:

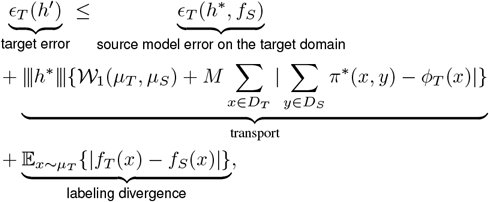

where 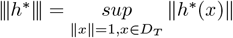.

## C Overview of methods for Transfer Learning and methods used in benchmarking

We provide here a short overview of standard methods used for Transfer Learning, with a focus on standard statistical, Machine Learning OT and non-OT-based methods. Standard statistical methods such as Correlation Alignment (*CA*) [Sun et al., 2017], Transfer Component Analysis (*TCA*) [Pan et al., 2010], Euclidean space data alignment [He and Wu, 2019] and Geodesic Flow Kernel (*GFK*) [Gong et al., 2012] tackle TL by aligning statistical properties between source and target domains, supporting both classification and regression tasks. In machine learning, OT-based TL has been, to the best of our knowledge, exclusively been studied for classification tasks and more specifically tested on image classification tasks, with methods such as Joint Distribution Adaptation [Long et al., 2013], Deep Joint Distribution Optimal Transport (*deepJDOT*) [Damodaran et al., 2018], and Class-aware Sample Reweighting [Wang et al., 2023]. These methods leverage OT to reduce the Wasserstein distance between source and target domains, thereby aligning their distributions. On the other hand, non-OT-based TL has been studied for classification including moment-matching methods[Long et al., 2015, 2017, Maria Carlucci et al., 2017] and adversarial learning methods [Ganin and Lempitsky, 2015, Tzeng et al., 2015, Ganin et al., 2016, Luo et al., 2017, Long et al., 2018, Zhang et al., 2019, Peng et al., 2019] as well as for regression such as Representation Subspace Distance (*RSD*) [Chen et al., 2021] and inverse GRAM matrices (*daregram*) [Nejjar et al., 2023], which learn a shared feature extractor by minimizing some discrepancies of source and target features.

## D Supplementary Figures

### D.1 Feature embedding differences

**Figure S1:**
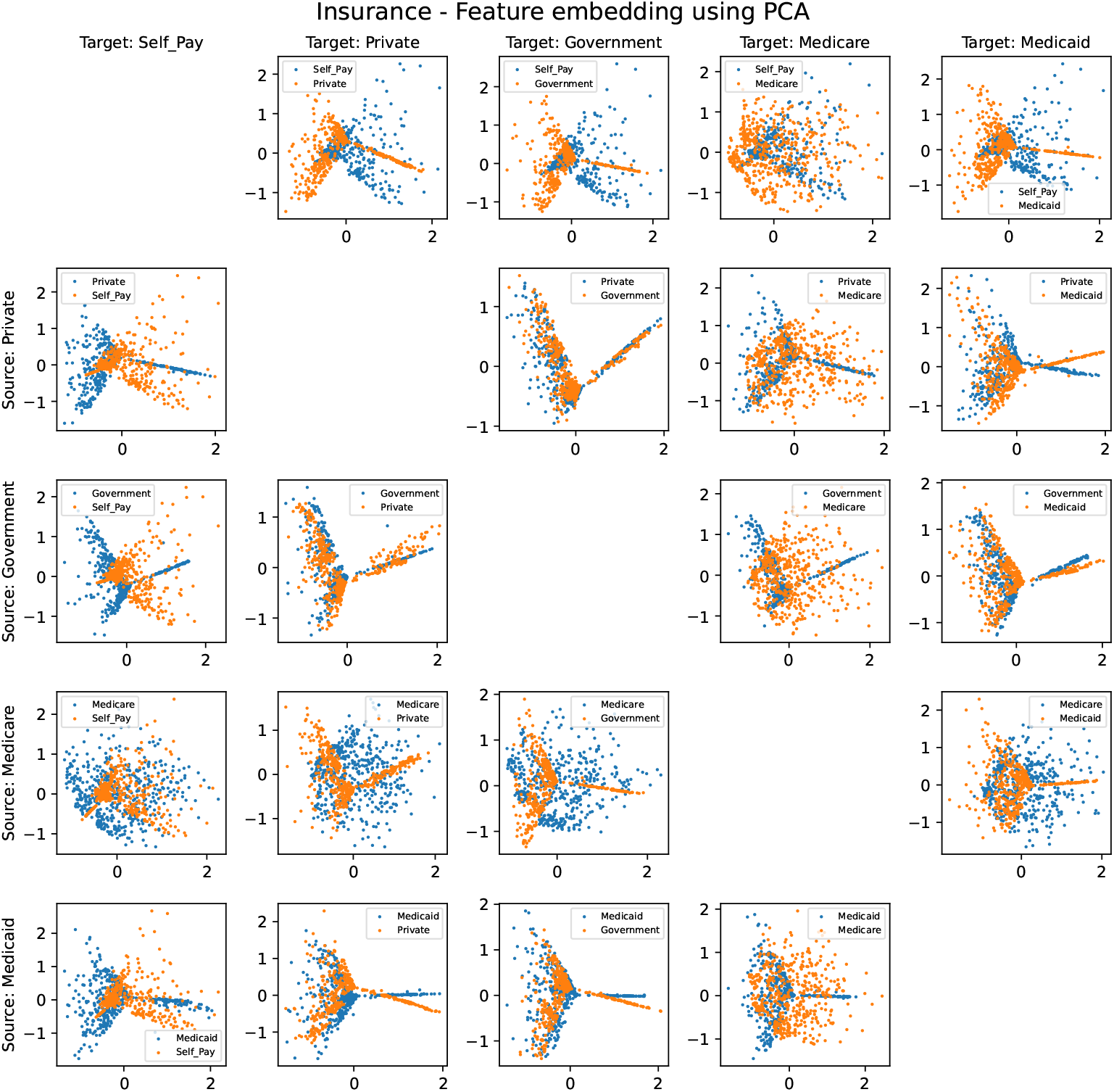
Feature embedding for pairwise insurance groups using PCA in the dimension of the first two principal components. Insurance groups include “Self_Pay,” “Private,” “Government,” “Medicare,” and “Medicaid.”

**Figure S2:**
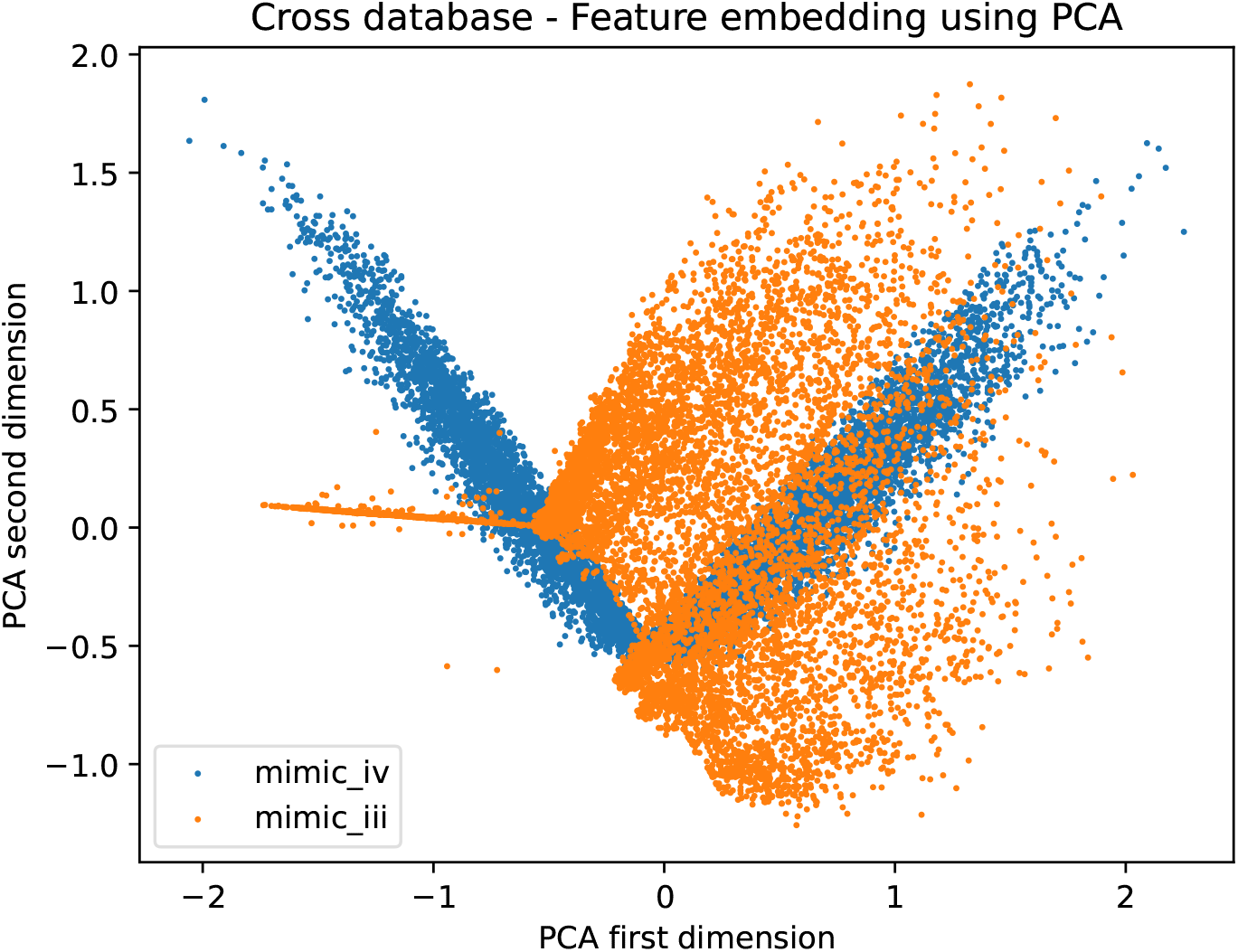
Feature embedding for MIMIC-IV (source) and MIMIC-III (target) using PCA in the dimension of the first two principal components.

Figure S3: Feature embedding for pairwise hospital groups using PCA in the dimension of the first two principal components. Hospital IDs include 420, 264, 243, 338, 73, 458, 167, 443, 208 and 300.

**Figure S4:**
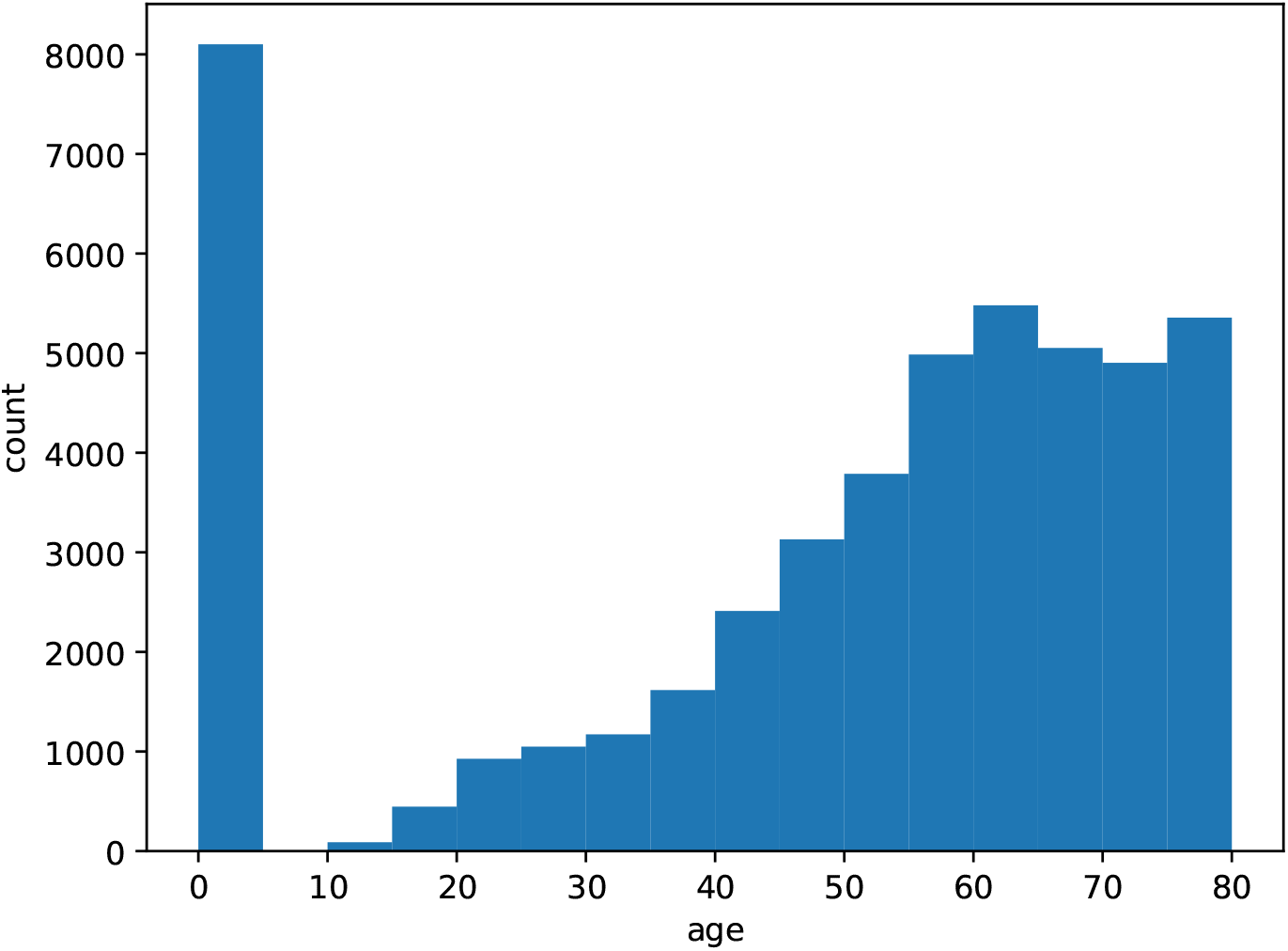
Age distribution for MIMIC-III.

### D.2 Illustration of *ϵ*_*T*_ (*h*\*, *f*_*S*_) in Theorem 1

**Figure S5:**
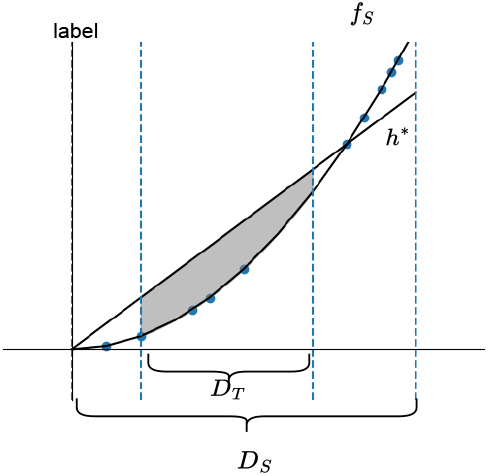
Illustration of *ϵ*_*T*_ (*h*\*, *f*_*S*_). *D*_*S*_ and *D*_*T*_ are source and target embedding spaces. The blue dots denote the source embeddings and source labels. *f*_*S*_ is the ground-truth labeling function for source embedding features and source labels. *h*\* is the source model trained by source embedding features and source labels. The gray area denotes *ϵ*_*T*_ (*h*\*, *f*_*S*_).

### D.3 Bound analysis

**Figure S6:**
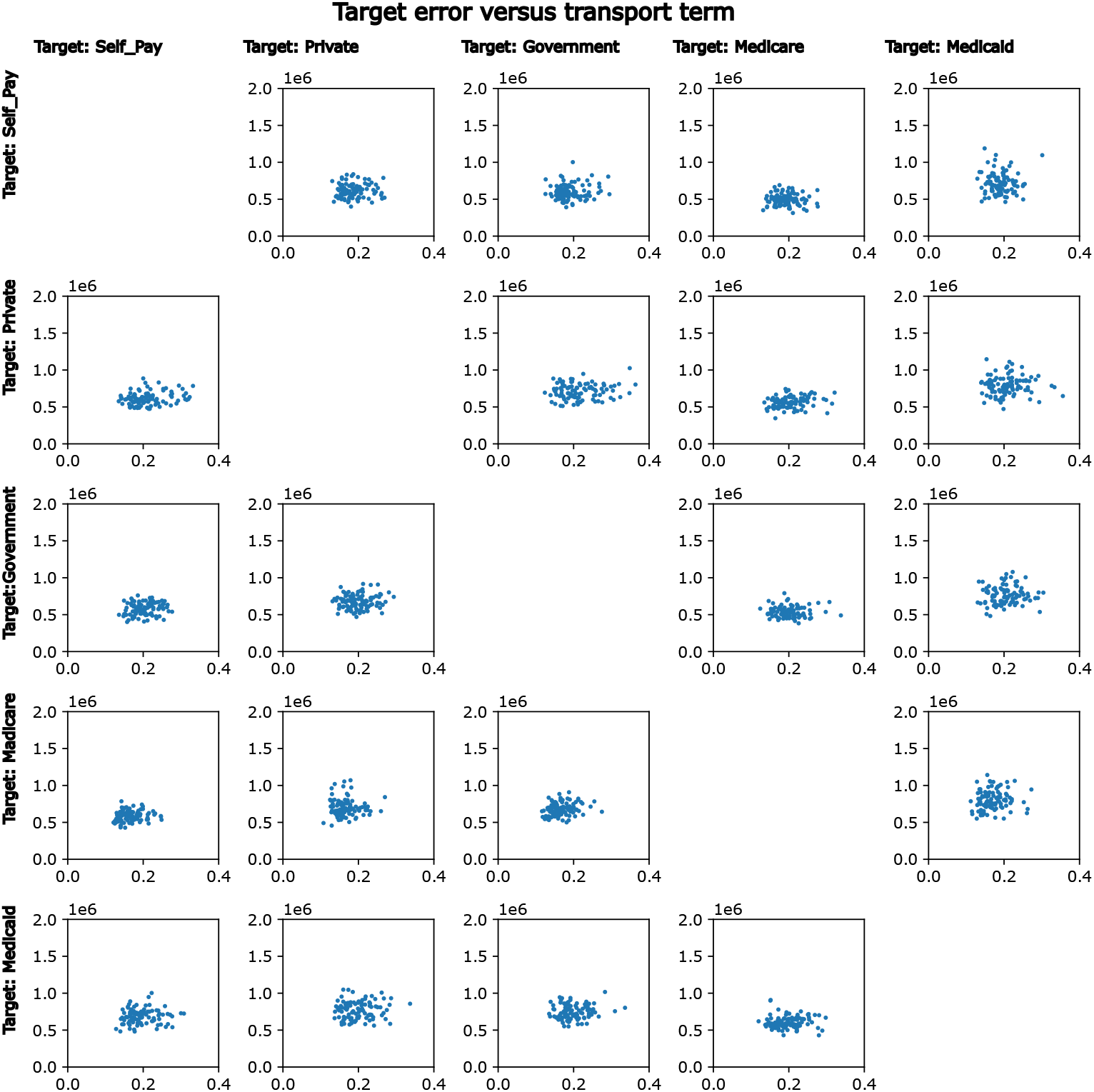
Bound analysis for pairwise insurance group experiments with respect to transport term. Target error versus transport term for pairwise insurance groups with an average PCC of 0.09.

**Figure S7:**
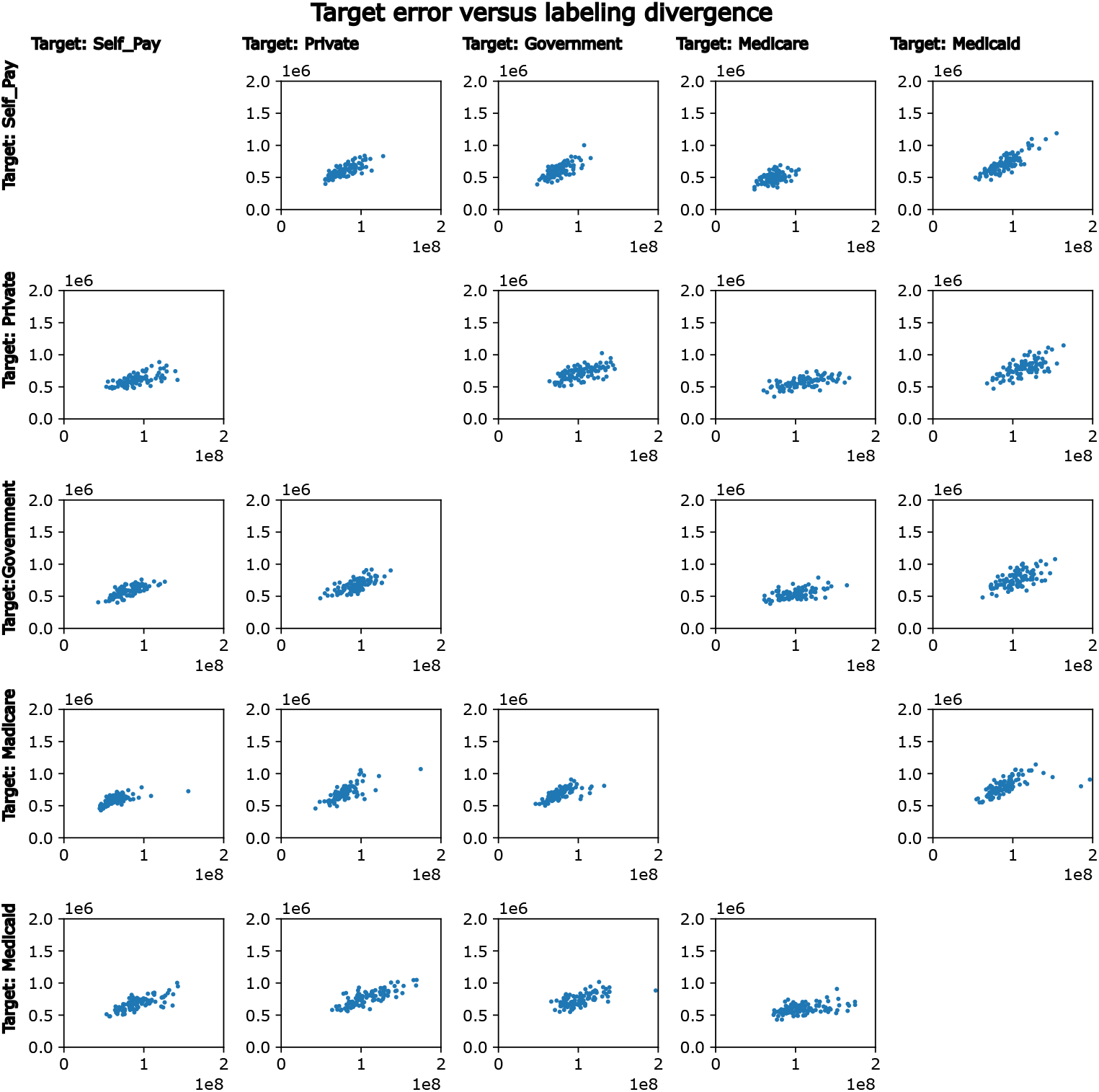
Bound analysis for pairwise insurance group experiments with respect to labeling divergence. Target error versus labeling divergence for pairwise insurance groups with an average PCC of 0.67.

### D.4 Admissions with significantly reduced durations in hospital for all pairwase experiments

**Figure S8:**
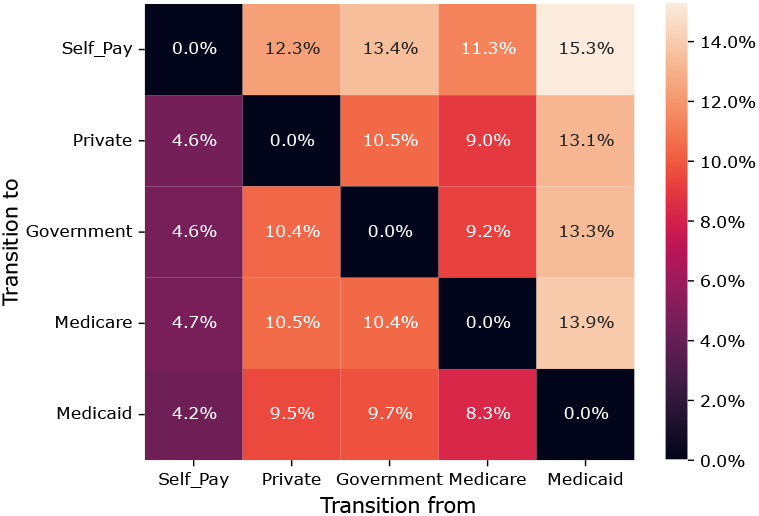
Percentages of admissions having significantly reduced durations in hospital for all pairwise insurance group experiments. For example, when transitioning to self paid insurance plans, 15.3% of the admissions on Medicaid would result in significantly reduced durations in hospital.

## E Supplementary Tables

### E.1 Mann-Whitney U test result for different experiments

**Table S1:** P-values from Mann-Whitney U tests to test the difference in distribution medians for the first two dimensions in PCA embeddings between pairwise insurance groups for insurance experiments. The significant p-values (*<* 0.05) are highlighted in red.

| Source | Target | dimension 1 | dimension 2 |
| --- | --- | --- | --- |
| Self_Pay | Private | 2.08e-02 | 3.74e-02 |
| Self_Pay | Government | 1.20e-01 | 5.41e-01 |
| Self_Pay | Medicare | 9.45e-02 | 3.74e-01 |
| Self_Pay | Medicaid | 1.79e-01 | 1.45e-01 |
| Private | Self_Pay | 2.03e-01 | 2.87e-03 |
| Private | Government | 4.81e-01 | 6.91e-01 |
| Private | Medicare | 1.16e-01 | 4.05e-01 |
| Private | Medicaid | 6.78e-01 | 9.21e-01 |
| Government | Self_Pay | 3.77e-01 | 1.54e-01 |
| Government | Private | 6.72e-01 | 2.91e-01 |
| Government | Medicare | 5.37e-03 | 6.38e-01 |
| Government | Medicaid | 8.21e-01 | 4.19e-01 |
| Medicare | Self_Pay | 6.89e-01 | 1.98e-01 |
| Medicare | Private | 8.58e-01 | 1.64e-01 |
| Medicare | Government | 2.88e-01 | 9.17e-01 |
| Medicare | Medicaid | 7.73e-01 | 6.56e-01 |
| Medicaid | Self_Pay | 1.46e-01 | 2.82e-02 |
| Medicaid | Private | 5.71e-02 | 4.99e-01 |
| Medicaid | Government | 3.10e-01 | 8.42e-01 |
| Medicaid | Medicare | 1.92e-02 | 2.96e-01 |

**Table S2:** P-values from Mann-Whitney U tests to test the difference in distribution medians for the first two dimensions in PCA embeddings between pairwise insurance groups for cross-database experiments. The significant p-values (*<* 0.05) are highlighted in red.

| Source | Target | dimension 1 | dimension 2 |
| --- | --- | --- | --- |
| MIMIC-IV | MIMIC-III | 5.39e-01 | 9.44e-08 |

**Table S3:** P-values from Mann-Whitney U tests to test the difference in distribution medians for the first two dimensions in PCA embeddings between pairwise hospitals for cross-hospital experiments. The significant p-values (<0.05) are highlighted in red.

| Source | Target | Dimension 1 | Dimension 2 |
| --- | --- | --- | --- |
| 420 | 264 | 1.86e-02 | 1.87e-02 |
| 420 | 243 | 2.06e-02 | 7.46e-01 |
| 420 | 338 | 1.54e-03 | 6.63e-02 |
| 420 | 73 | 1.13e-01 | 5.40e-01 |
| 420 | 458 | 4.80e-02 | 6.11e-01 |
| 420 | 167 | 2.16e-02 | 1.88e-01 |
| 420 | 443 | 2.23e-04 | 2.97e-01 |
| 420 | 208 | 2.06e-03 | 6.32e-01 |
| 420 | 300 | 2.27e-04 | 1.23e-01 |
| 264 | 420 | 1.47e-03 | 1.09e-01 |
| 264 | 243 | 1.24e-05 | 1.51e-03 |
| 264 | 338 | 4.91e-06 | 2.08e-03 |
| 264 | 73 | 5.04e-05 | 4.79e-10 |
| 264 | 458 | 1.06e-15 | 2.61e-15 |
| 264 | 167 | 6.50e-12 | 2.32e-02 |
| 264 | 443 | 1.49e-01 | 2.41e-08 |
| 264 | 208 | 1.23e-10 | 3.96e-03 |
| 264 | 300 | 4.02e-01 | 8.48e-23 |
| 243 | 420 | 7.98e-03 | 8.67e-01 |
| 243 | 264 | 1.09e-04 | 2.42e-01 |
| 243 | 338 | 1.48e-03 | 3.93e-12 |
| 243 | 73 | 3.88e-01 | 5.64e-01 |
| 243 | 458 | 1.48e-03 | 2.31e-09 |
| 243 | 167 | 5.80e-02 | 8.58e-02 |
| 243 | 443 | 5.84e-04 | 8.12e-02 |
| 243 | 208 | 1.34e-01 | 1.54e-10 |
| 243 | 300 | 7.24e-11 | 8.64e-08 |
| 338 | 420 | 2.72e-04 | 9.26e-01 |
| 338 | 264 | 1.10e-10 | 1.44e-07 |
| 338 | 243 | 3.84e-05 | 7.09e-02 |
| 338 | 73 | 2.38e-05 | 6.55e-06 |
| 338 | 458 | 2.92e-13 | 6.63e-14 |
| 338 | 167 | 5.61e-13 | 1.54e-16 |
| 338 | 443 | 3.36e-02 | 2.03e-02 |
| 338 | 208 | 4.51e-20 | 9.30e-01 |
| 338 | 300 | 4.44e-06 | 5.64e-13 |
| 73 | 420 | 5.28e-02 | 5.25e-01 |
| 73 | 264 | 8.80e-05 | 4.75e-03 |
| 73 | 243 | 1.48e-01 | 4.99e-01 |
| 73 | 338 | 3.55e-04 | 1.59e-02 |
| 73 | 458 | 1.71e-03 | 6.37e-02 |
| 73 | 167 | 6.12e-03 | 1.38e-03 |
| 73 | 443 | 2.84e-03 | 9.99e-06 |
| 73 | 208 | 8.45e-03 | 7.82e-02 |
| 73 | 300 | 1.10e-05 | 3.70e-08 |
| 458 | 243 | 1.48e-03 | 4.06e-02 |
| 458 | 338 | 8.57e-19 | 1.04e-07 |
| 458 | 73 | 3.40e-02 | 4.80e-03 |
| 458 | 167 | 1.36e-07 | 5.00e-06 |
| 458 | 443 | 1.36e-14 | 4.05e-14 |
| 458 | 208 | 4.62e-11 | 5.51e-24 |
| 458 | 300 | 7.81e-20 | 7.75e-03 |
| 167 | 420 | 2.06e-02 | 1.42e-01 |
| 167 | 264 | 4.18e-12 | 1.59e-06 |
| 167 | 243 | 7.08e-01 | 2.09e-04 |
| 167 | 338 | 3.75e-14 | 4.52e-13 |
| 167 | 73 | 1.75e-01 | 1.33e-01 |
| 167 | 458 | 4.11e-05 | 1.65e-01 |
| 167 | 443 | 5.51e-13 | 6.42e-05 |
| 167 | 208 | 7.90e-18 | 4.13e-05 |
| 167 | 300 | 3.88e-15 | 1.64e-04 |
| 443 | 420 | 4.04e-03 | 5.46e-01 |
| 443 | 264 | 3.08e-01 | 7.18e-10 |
| 443 | 243 | 6.07e-04 | 6.30e-03 |
| 443 | 338 | 5.02e-18 | 6.87e-14 |
| 443 | 73 | 1.27e-05 | 1.41e-03 |
| 443 | 458 | 3.35e-13 | 5.57e-02 |
| 443 | 167 | 3.14e-12 | 3.07e-23 |
| 443 | 208 | 3.11e-05 | 8.78e-02 |
| 443 | 300 | 4.97e-09 | 3.99e-40 |
| 208 | 420 | 1.04e-02 | 2.39e-01 |
| 208 | 264 | 3.00e-09 | 7.67e-08 |
| 208 | 243 | 3.33e-08 | 1.20e-03 |
| 208 | 338 | 4.54e-08 | 3.52e-20 |
| 208 | 73 | 4.04e-04 | 4.14e-03 |
| 208 | 458 | 3.65e-11 | 5.54e-10 |
| 208 | 167 | 2.71e-12 | 2.24e-12 |
| 208 | 443 | 7.28e-01 | 3.54e-09 |
| 208 | 300 | 4.98e-26 | 3.69e-18 |
| 300 | 420 | 1.75e-03 | 3.32e-01 |
| 300 | 264 | 1.72e-15 | 6.31e-01 |
| 300 | 243 | 2.68e-05 | 3.22e-14 |
| 300 | 338 | 1.91e-02 | 2.60e-07 |
| 300 | 73 | 1.94e-04 | 5.59e-05 |
| 300 | 458 | 9.68e-20 | 9.01e-05 |
| 300 | 167 | 4.82e-16 | 4.25e-01 |
| 300 | 443 | 3.96e-17 | 1.52e-11 |
| 300 | 208 | 7.16e-15 | 4.63e-14 |

### E.2 Benchmark results for pairwise insurance group experiments

**Table S4:** Benchmark results for pairwise insurance group experiments. Medians and standard deviations of the log of *MAE* between projected duration and observed duration on target admissions for different insurance groups using *OTTEHR, TCA, CA, GFK, deepJDOT, RSD* and *daregram*. The outperformance ratio of *OTTEHR* to *TCA*/*CA*/*GFK*/*RSD*/*daregram* is defined as the percentage decrease in the median *MAE* from *TCA*/*CA*/*GFK*/*RSD*/*daregram* to *OTTEHR*. The outperformance ratio of *OTTEHR* to *deepJDOT* is defined as the percentage decrease in median of the log-transformed of standard deviation of *MAE* from *deepJDOT* to *OTTEHR*.

| Source | Target | *OTTEHR* | *TCA* | *CA* | *GFK* | *deepJDOT* | *RSD* | *daregram* |
| --- | --- | --- | --- | --- | --- | --- | --- | --- |
| Self_Pay | Private | 13.40(0.16) | 13.60(0.17) | 13.63(0.18) | 13.60(0.17) | 13.32(0.18) | 13.68(0.13) | 13.66(0.14) |
| Self_Pay | Government | 13.35(0.15) | 13.55(0.16) | 13.56(0.17) | 13.56(0.16) | 13.32(0.17) | 13.64(0.12) | 13.62(0.13) |
| Self_Pay | Medicare | 13.28(0.14) | 13.54(0.14) | 13.58(0.15) | 13.55(0.15) | 13.13(0.47) | 13.64(0.11) | 13.67(0.10) |
| Self_Pay | Medicaid | 13.53(0.18) | 13.74(0.18) | 13.73(0.18) | 13.73(0.18) | 13.46(0.29) | 13.79(0.14) | 13.78(0.12) |
| Private | Self_Pay | 13.36(0.17) | 13.78(0.19) | 13.80(0.17) | 13.78(0.20) | 13.30(0.87) | 13.21(0.13) | 13.23(0.10) |
| Private | Government | 13.52(0.16) | 13.83(0.18) | 13.86(0.19) | 13.84(0.18) | 13.44(0.31) | 13.62(0.14) | 13.61(0.11) |
| Private | Medicare | 13.48(0.16) | 13.95(0.21) | 13.94(0.21) | 13.94(0.21) | 13.19(0.13) | 13.63(0.10) | 13.66(0.10) |
| Private | Medicaid | 13.65(0.18) | 13.98(0.17) | 13.99(0.17) | 13.98(0.17) | 13.55(0.77) | 13.82(0.13) | 13.81(0.14) |
| Government | Self_Pay | 13.28(0.14) | 13.66(0.18) | 13.69(0.18) | 13.67(0.18) | 13.25(0.12) | 13.22(0.09) | 13.25(0.12) |
| Government | Private | 13.45(0.16) | 13.78(0.17) | 13.82(0.18) | 13.78(0.17) | 13.49(1.01) | 13.66(0.14) | 13.67(0.12) |
| Government | Medicare | 13.42(0.15) | 13.85(0.19) | 13.84(0.19) | 13.85(0.19) | 13.17(0.80) | 13.66(0.10) | 13.66(0.09) |
| Government | Medicaid | 13.62(0.16) | 13.91(0.16) | 13.93(0.17) | 13.92(0.16) | 13.56(0.55) | 13.81(0.13) | 13.82(0.14) |
| Medicare | Self_Pay | 13.23(0.13) | 13.44(0.18) | 13.47(0.18) | 13.44(0.18) | 13.28(0.81) | 13.22(0.12) | 13.22(0.12) |
| Medicare | Private | 13.44(0.17) | 13.62(0.17) | 13.65(0.18) | 13.62(0.17) | 13.45(1.06) | 13.67(0.14) | 13.68(0.12) |
| Medicare | Government | 13.42(0.13) | 13.58(0.17) | 13.62(0.15) | 13.57(0.17) | 13.43(0.36) | 13.64(0.13) | 13.64(0.12) |
| Medicare | Medicaid | 13.61(0.17) | 13.74(0.20) | 13.78(0.20) | 13.74(0.20) | 13.55(0.22) | 13.81(0.11) | 13.77(0.14) |
| Medicaid | Self_Pay | 13.42(0.18) | 13.77(0.19) | 13.81(0.20) | 13.78(0.19) | 13.42(0.87) | 13.23(0.12) | 13.22(0.11) |
| Medicaid | Private | 13.64(0.18) | 13.88(0.20) | 13.95(0.19) | 13.87(0.20) | 13.54(0.17) | 13.66(0.16) | 13.66(0.13) |
| Medicaid | Government | 13.55(0.16) | 13.87(0.17) | 13.89(0.16) | 13.87(0.18) | 13.53(0.81) | 13.63(0.13) | 13.66(0.13) |
| Medicaid | Medicare | 13.54(0.16) | 13.92(0.21) | 13.96(0.21) | 13.92(0.21) | 13.32(0.25) | 13.66(0.10) | 13.65(0.10) |
| Population Median |  | 13.46(0.20) | 13.75(0.23) | 13.78(0.23) | 13.75(0.23) | 13.39(0.62) | 13.64(0.24) | 13.64(0.23) |
| Outperformance ratio of *OTTEHR* |  | N.A. | 25.17 | 27.39 | 25.17 | 67.74 | 16.47 | 16.47 |

**Table S5:**
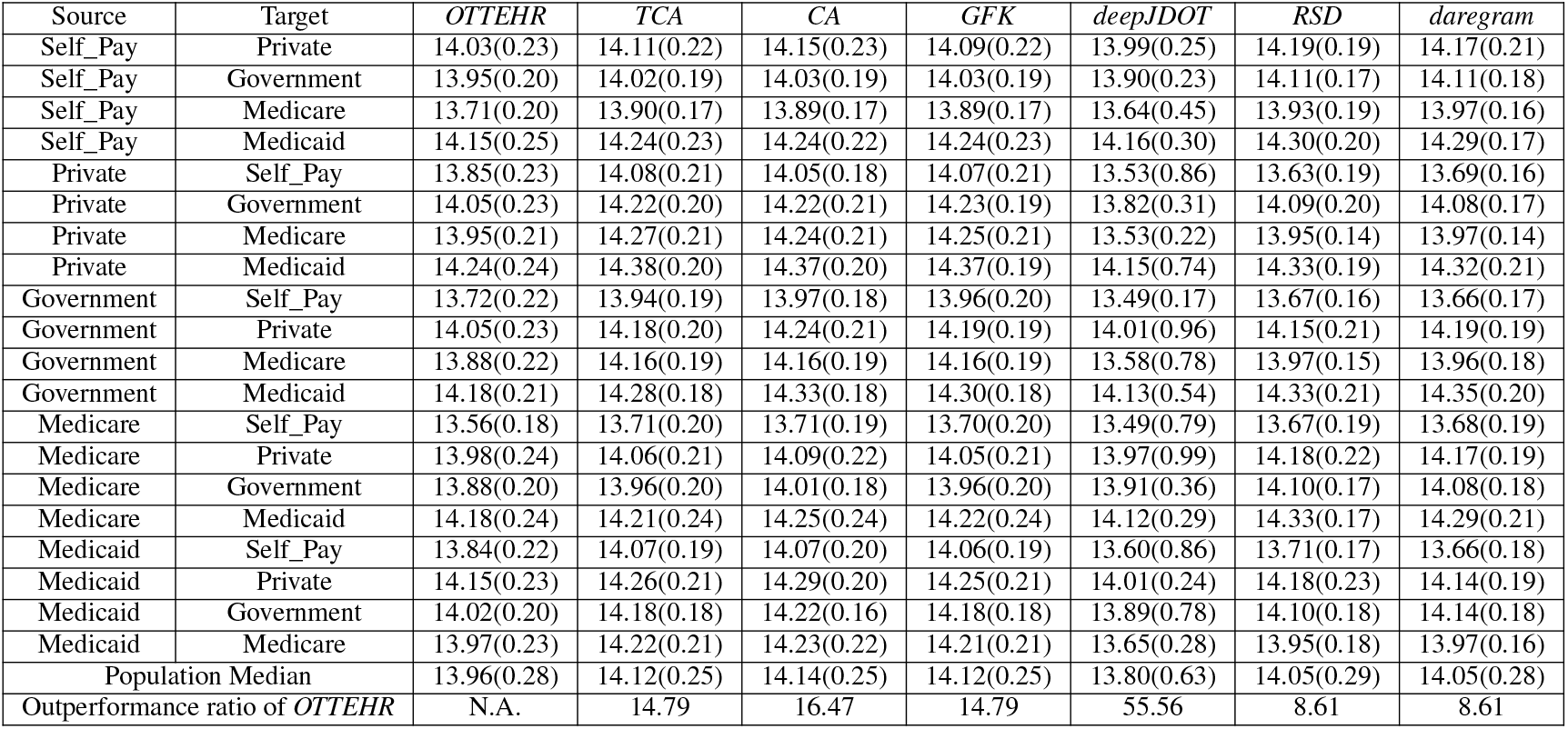
Benchmark results for pairwise insurance group experiments. Medians and standard deviations of the log of *RMSE* between projected duration and observed duration on target admissions for different insurance groups using *OTTEHR, TCA, CA, GFK, deepJDOT, RSD* and *daregram*. The outperformance ratio of *OTTEHR* to *TCA*/*CA*/*GFK*/*RSD*/*daregram* is defined as the percentage decrease in the median *RMSE* from *TCA*/*CA*/*GFK*/*RSD*/*daregram* to *OTTEHR*. The outperformance ratio of *OTTEHR* to *deepJDOT* is defined as the percentage decrease in median of the log-transformed of standard deviation of *RMSE* from *deepJDOT* to *OTTEHR*.

**Table S6:**
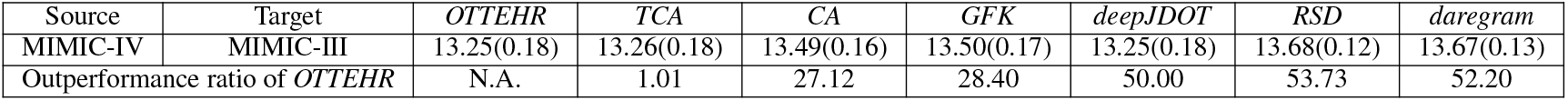
Benchmark results for cross-database experiments. Medians and standard deviations of the log of *MAE* between projected duration and observed duration on target admissions for the cross-database experiment using *OTTEHR, TCA, CA, GFK, deepJDOT, RSD* and *daregram*. The outperformance ratio of *OTTEHR* to *TCA*/*CA*/*GFK*/*RSD*/*daregram* is defined as the percentage decrease in the median *MAE* from *TCA*/*CA*/*GFK*/*RSD*/*daregram* to *OTTEHR*. The outperformance ratio of *OTTEHR* to *deepJDOT* is defined as the percentage decrease in median of the log-transformed of standard deviation of *MAE* from *deepJDOT* to *OTTEHR*.

| Source | Target | *OTTEHR* | *TCA* | *CA* | *GFK* | *deepJDOT* | *RSD* | *daregram* |
| --- | --- | --- | --- | --- | --- | --- | --- | --- |
| MIMIC-IV | MIMIC-III | 13.25(0.18) | 13.26(0.18) | 13.49(0.16) | 13.50(0.17) | 13.25(0.18) | 13.68(0.12) | 13.67(0.13) |
| Outperformance ratio of *OTTEHR* |  | N.A. | 1.01 | 27.12 | 28.40 | 50.00 | 53.73 | 52.20 |

**Table S7:** Benchmark results for cross-database experiments. Medians and standard deviations of the log of *RMSE* between projected duration and observed duration on target admissions for the cross-database experiment using *OTTEHR, TCA, CA, GFK, deepJDOT, RSD* and *daregram*. The outperformance ratio of *OTTEHR* to *TCA*/*CA*/*GFK*/*RSD*/*daregram* is defined as the percentage decrease in the median *RMSE* from *TCA*/*CA*/*GFK*/*RSD*/*daregram* to *OTTEHR*. The outperformance ratio of *OTTEHR* to *deepJDOT* is defined as the percentage decrease in median of the log-transformed of standard deviation of *RMSE* from *deepJDOT* to *OTTEHR*.

| Source | Target | *OTTEHR* | *TCA* | *CA* | *GFK* | *deepJDOT* | *RSD* | *daregram* |
| --- | --- | --- | --- | --- | --- | --- | --- | --- |
| MIMIC-IV | MIMIC-III | 13.90(0.30) | 13.94(0.29) | 14.01(0.24) | 14.01(0.24) | 13.90(0.41) | 14.08(0.20) | 14.05(0.23) |
| Outperformance ratio of *OTTEHR* |  | N.A. | 3.92 | 11.17 | 11.17 | 36.67 | 18.95 | 15.55 |

**Table S8:** Benchmark results for pairwise cross-hospital experiments. Medians and standard deviations of the log of *MAE* between projected duration and observed duration on target admissions for different insurance groups using *OTTEHR, TCA, CA, GFK, deepJDOT, RSD* and *daregram*. The outperformance ratio of *OTTEHR* to *TCA*/*CA*/*GFK*/*RSD*/*daregram* is defined as the percentage decrease in the median *MAE* from *TCA*/*CA*/*GFK*/*RSD*/*daregram* to *OTTEHR*. The outperformance ratio of *OTTEHR* to *deepJDOT* is defined as the percentage decrease in median of the log-transformed of standard deviation of *MAE* from *deepJDOT* to *OTTEHR*.

| Source | Target | *OTTEHR* | *TCA* | *CA* | *GFK* | *deepJDOT* | *RSD* | *daregram* |
| --- | --- | --- | --- | --- | --- | --- | --- | --- |
| 420 | 264 | 13.07(0.13) | 13.20(0.12) | 13.29(0.13) | 13.27(0.12) | 19.02(1.91) | 13.41(0.08) | 13.42(0.09) |
| 420 | 243 | 13.08(0.15) | 13.19(0.12) | 13.38(0.15) | 13.28(0.13) | 18.80(1.92) | 13.40(0.10) | 13.42(0.11) |
| 420 | 338 | 13.11(0.11) | 13.22(0.10) | 13.32(0.11) | 13.28(0.10) | 18.60(1.72) | 13.49(0.09) | 13.50(0.10) |
| 420 | 73 | 13.37(0.16) | 13.37(0.15) | 13.52(0.16) | 13.46(0.15) | 18.64(1.76) | 13.74(0.12) | 13.76(0.11) |
| 420 | 458 | 13.42(0.29) | 13.48(0.27) | 13.57(0.25) | 13.52(0.26) | 18.71(1.34) | 13.76(0.21) | 13.75(0.22) |
| 420 | 167 | 13.29(0.14) | 13.34(0.12) | 13.47(0.15) | 13.38(0.13) | 18.43(1.33) | 13.67(0.10) | 13.64(0.12) |
| 420 | 443 | 13.52(0.14) | 13.53(0.13) | 13.58(0.13) | 13.58(0.13) | 18.87(1.49) | 13.87(0.12) | 13.87(0.11) |
| 420 | 208 | 13.30(0.16) | 13.35(0.16) | 13.45(0.16) | 13.41(0.15) | 18.80(1.97) | 13.65(0.12) | 13.67(0.15) |
| 420 | 300 | 13.17(0.15) | 13.29(0.13) | 13.35(0.14) | 13.33(0.14) | 18.91(1.80) | 13.54(0.11) | 13.54(0.11) |
| 264 | 420 | 13.38(0.13) | 13.52(0.19) | 13.99(0.19) | 13.85(0.15) | 13.64(1.99) | 13.68(0.10) | 13.69(0.11) |
| 264 | 243 | 13.10(0.13) | 13.13(0.16) | 13.48(0.20) | 13.34(0.16) | 14.05(1.87) | 13.42(0.11) | 13.43(0.10) |
| 264 | 338 | 13.13(0.12) | 13.12(0.13) | 13.35(0.17) | 13.25(0.15) | 14.23(1.37) | 13.49(0.09) | 13.49(0.08) |
| 264 | 73 | 13.39(0.14) | 13.40(0.16) | 13.71(0.16) | 13.58(0.15) | 14.25(1.20) | 13.76(0.11) | 13.74(0.10) |
| 264 | 458 | 13.43(0.27) | 13.42(0.27) | 13.65(0.25) | 13.57(0.26) | 14.15(2.10) | 13.75(0.23) | 13.70(0.18) |
| 264 | 167 | 13.30(0.13) | 13.33(0.13) | 13.57(0.16) | 13.46(0.13) | 14.52(2.95) | 13.65(0.11) | 13.65(0.11) |
| 264 | 443 | 13.55(0.14) | 13.53(0.14) | 13.70(0.15) | 13.65(0.13) | 14.15(1.73) | 13.87(0.12) | 13.87(0.12) |
| 264 | 208 | 13.30(0.17) | 13.29(0.17) | 13.47(0.16) | 13.42(0.16) | 14.20(1.33) | 13.63(0.14) | 13.64(0.14) |
| 264 | 300 | 13.21(0.14) | 13.19(0.14) | 13.43(0.15) | 13.34(0.12) | 14.12(2.02) | 13.53(0.11) | 13.54(0.10) |
| 243 | 420 | 13.39(0.13) | 13.44(0.18) | 13.87(0.22) | 13.77(0.18) | 14.75(1.95) | 13.70(0.11) | 13.71(0.11) |
| 243 | 264 | 13.03(0.11) | 13.04(0.10) | 13.29(0.16) | 13.22(0.13) | 15.07(1.43) | 13.40(0.10) | 13.41(0.11) |
| 243 | 338 | 13.06(0.13) | 13.08(0.12) | 13.27(0.17) | 13.23(0.16) | 15.01(2.02) | 13.47(0.09) | 13.52(0.10) |
| 243 | 73 | 13.38(0.15) | 13.37(0.16) | 13.60(0.16) | 13.53(0.15) | 15.15(1.90) | 13.76(0.11) | 13.74(0.12) |
| 243 | 458 | 13.42(0.33) | 13.40(0.33) | 13.56(0.29) | 13.48(0.31) | 15.18(2.64) | 13.75(0.20) | 13.78(0.20) |
| 243 | 167 | 13.27(0.15) | 13.25(0.14) | 13.52(0.16) | 13.39(0.14) | 15.20(1.59) | 13.63(0.11) | 13.66(0.10) |
| 243 | 443 | 13.52(0.14) | 13.47(0.14) | 13.63(0.13) | 13.60(0.13) | 15.04(1.71) | 13.86(0.11) | 13.86(0.12) |
| 243 | 208 | 13.28(0.17) | 13.27(0.17) | 13.46(0.17) | 13.41(0.17) | 15.07(1.73) | 13.65(0.11) | 13.65(0.13) |
| 243 | 300 | 13.14(0.15) | 13.15(0.14) | 13.33(0.17) | 13.27(0.14) | 15.09(1.15) | 13.53(0.12) | 13.55(0.12) |
| 338 | 420 | 13.41(0.12) | 13.55(0.17) | 14.09(0.22) | 13.94(0.14) | 13.74(1.68) | 13.70(0.10) | 13.69(0.10) |
| 338 | 264 | 13.13(0.11) | 13.13(0.11) | 13.41(0.17) | 13.31(0.11) | 14.25(2.34) | 13.42(0.08) | 13.41(0.10) |
| 338 | 243 | 13.15(0.13) | 13.16(0.14) | 13.55(0.19) | 13.41(0.15) | 13.84(1.35) | 13.41(0.10) | 13.39(0.10) |
| 338 | 73 | 13.41(0.13) | 13.42(0.14) | 13.76(0.17) | 13.64(0.13) | 14.11(1.97) | 13.74(0.11) | 13.72(0.12) |
| 338 | 458 | 13.47(0.26) | 13.47(0.26) | 13.73(0.26) | 13.61(0.25) | 14.31(2.11) | 13.77(0.20) | 13.76(0.16) |
| 338 | 167 | 13.33(0.13) | 13.32(0.13) | 13.59(0.18) | 13.47(0.14) | 14.31(1.92) | 13.67(0.11) | 13.63(0.11) |
| 338 | 443 | 13.55(0.15) | 13.53(0.15) | 13.76(0.18) | 13.67(0.15) | 14.29(1.85) | 13.86(0.11) | 13.87(0.09) |
| 338 | 208 | 13.30(0.14) | 13.31(0.15) | 13.57(0.18) | 13.48(0.15) | 14.17(1.19) | 13.65(0.13) | 13.66(0.13) |
| 338 | 300 | 13.24(0.13) | 13.21(0.14) | 13.48(0.16) | 13.38(0.14) | 14.08(2.59) | 13.54(0.11) | 13.55(0.10) |
| 73 | 420 | 13.50(0.14) | 13.58(0.17) | 14.11(0.20) | 13.96(0.20) | 16.32(2.36) | 13.71(0.10) | 13.72(0.11) |
| 73 | 264 | 13.17(0.12) | 13.27(0.12) | 13.50(0.15) | 13.42(0.16) | 16.84(2.10) | 13.41(0.08) | 13.39(0.10) |
| 73 | 243 | 13.22(0.15) | 13.29(0.15) | 13.71(0.20) | 13.54(0.17) | 16.30(2.32) | 13.44(0.09) | 13.41(0.10) |
| 73 | 338 | 13.20(0.13) | 13.29(0.12) | 13.54(0.17) | 13.46(0.16) | 16.67(1.93) | 13.50(0.10) | 13.50(0.10) |
| 73 | 458 | 13.48(0.28) | 13.50(0.26) | 13.78(0.24) | 13.67(0.25) | 16.47(2.92) | 13.75(0.19) | 13.75(0.21) |
| 73 | 167 | 13.36(0.13) | 13.40(0.13) | 13.66(0.18) | 13.57(0.15) | 16.60(1.91) | 13.67(0.10) | 13.68(0.10) |
| 73 | 443 | 13.56(0.14) | 13.57(0.14) | 13.80(0.14) | 13.73(0.14) | 16.51(2.00) | 13.86(0.10) | 13.85(0.10) |
| 73 | 208 | 13.35(0.14) | 13.44(0.14) | 13.67(0.17) | 13.57(0.15) | 17.01(2.09) | 13.65(0.11) | 13.65(0.13) |
| 73 | 300 | 13.23(0.15) | 13.32(0.13) | 13.56(0.15) | 13.50(0.14) | 16.88(1.77) | 13.53(0.12) | 13.54(0.12) |
| 458 | 420 | 13.64(0.43) | 13.74(0.51) | 14.47(0.67) | 14.27(0.71) | 15.40(2.87) | 13.70(0.09) | 13.70(0.10) |
| 458 | 264 | 13.31(0.25) | 13.34(0.27) | 13.76(0.53) | 13.65(0.55) | 16.31(3.37) | 13.42(0.10) | 13.41(0.10) |
| 458 | 243 | 13.36(0.32) | 13.35(0.37) | 13.96(0.59) | 13.78(0.63) | 15.89(1.96) | 13.41(0.08) | 13.42(0.11) |
| 458 | 338 | 13.31(0.23) | 13.34(0.27) | 13.75(0.50) | 13.65(0.54) | 15.66(2.03) | 13.49(0.09) | 13.48(0.09) |
| 458 | 73 | 13.58(0.30) | 13.55(0.36) | 14.13(0.58) | 13.93(0.58) | 15.68(2.73) | 13.75(0.12) | 13.77(0.13) |
| 458 | 167 | 13.44(0.24) | 13.47(0.23) | 13.89(0.49) | 13.70(0.52) | 15.87(1.70) | 13.64(0.12) | 13.64(0.10) |
| 458 | 443 | 13.64(0.19) | 13.63(0.19) | 14.04(0.48) | 13.91(0.49) | 15.82(2.10) | 13.85(0.10) | 13.88(0.11) |
| 458 | 208 | 13.44(0.23) | 13.43(0.25) | 13.84(0.47) | 13.68(0.50) | 16.03(2.06) | 13.65(0.13) | 13.64(0.12) |
| 458 | 300 | 13.37(0.21) | 13.41(0.24) | 13.84(0.50) | 13.71(0.51) | 15.64(2.34) | 13.54(0.13) | 13.53(0.12) |

**Table S9:** Benchmark results for pairwise cross-hospital experiments. Medians and standard deviations of the log of *MAE* between projected duration and observed duration on target admissions for different insurance groups using *OTTEHR, TCA, CA, GFK, deepJDOT, RSD* and *daregram*. The outperformance ratio of *OTTEHR* to *TCA*/*CA*/*GFK*/*RSD*/*daregram* is defined as the percentage decrease in the median *MAE* from *TCA*/*CA*/*GFK*/*RSD*/*daregram* to *OTTEHR*. The outperformance ratio of *OTTEHR* to *deepJDOT* is defined as the percentage decrease in median of the log-transformed of standard deviation of *MAE* from *deepJDOT* to *OTTEHR*.

| Source | Target | *OTTEHR* | *TCA* | *CA* | *GFK* | *deepJDOT* | *RSD* | *daregram* |
| --- | --- | --- | --- | --- | --- | --- | --- | --- |
| 167 | 420 | 13.52(0.15) | 13.60(0.26) | 14.17(0.22) | 14.01(0.21) | 15.00(2.18) | 13.71(0.12) | 13.69(0.10) |
| 167 | 264 | 13.20(0.11) | 13.23(0.11) | 13.54(0.15) | 13.46(0.13) | 15.83(2.32) | 13.41(0.10) | 13.41(0.10) |
| 167 | 243 | 13.22(0.13) | 13.23(0.14) | 13.67(0.20) | 13.53(0.16) | 15.35(1.78) | 13.43(0.11) | 13.45(0.09) |
| 167 | 338 | 13.23(0.12) | 13.24(0.11) | 13.50(0.17) | 13.41(0.13) | 15.53(2.81) | 13.49(0.11) | 13.50(0.09) |
| 167 | 73 | 13.44(0.13) | 13.45(0.14) | 13.81(0.17) | 13.68(0.15) | 15.57(1.45) | 13.74(0.11) | 13.74(0.11) |
| 167 | 458 | 13.47(0.26) | 13.48(0.26) | 13.78(0.24) | 13.64(0.23) | 15.58(2.84) | 13.79(0.23) | 13.79(0.21) |
| 167 | 443 | 13.56(0.14) | 13.55(0.13) | 13.76(0.13) | 13.72(0.13) | 15.50(2.55) | 13.88(0.13) | 13.85(0.14) |
| 167 | 208 | 13.33(0.16) | 13.35(0.16) | 13.65(0.19) | 13.53(0.17) | 15.76(1.76) | 13.65(0.11) | 13.68(0.12) |
| 167 | 300 | 13.31(0.14) | 13.31(0.14) | 13.57(0.16) | 13.51(0.15) | 15.65(1.79) | 13.53(0.14) | 13.53(0.14) |
| 443 | 420 | 13.66(0.15) | 13.76(0.19) | 14.50(0.23) | 14.34(0.21) | 14.83(1.28) | 13.72(0.11) | 13.72(0.11) |
| 443 | 264 | 13.41(0.12) | 13.44(0.14) | 13.82(0.18) | 13.70(0.16) | 15.63(1.77) | 13.41(0.10) | 13.41(0.11) |
| 443 | 243 | 13.47(0.13) | 13.49(0.15) | 14.00(0.21) | 13.83(0.19) | 15.75(3.09) | 13.41(0.11) | 13.42(0.09) |
| 443 | 338 | 13.39(0.14) | 13.41(0.15) | 13.77(0.19) | 13.69(0.18) | 15.79(1.35) | 13.50(0.09) | 13.49(0.10) |
| 443 | 73 | 13.59(0.14) | 13.60(0.16) | 14.12(0.22) | 13.98(0.20) | 15.86(2.41) | 13.74(0.12) | 13.73(0.12) |
| 443 | 458 | 13.63(0.24) | 13.65(0.23) | 14.01(0.23) | 13.89(0.22) | 15.55(1.15) | 13.78(0.19) | 13.79(0.20) |
| 443 | 167 | 13.53(0.13) | 13.54(0.15) | 13.93(0.20) | 13.76(0.17) | 15.65(1.51) | 13.67(0.12) | 13.66(0.11) |
| 443 | 208 | 13.52(0.13) | 13.50(0.13) | 13.82(0.16) | 13.75(0.15) | 15.89(1.23) | 13.65(0.12) | 13.66(0.11) |
| 443 | 300 | 13.45(0.13) | 13.43(0.14) | 13.79(0.17) | 13.70(0.15) | 15.71(2.32) | 13.54(0.11) | 13.53(0.12) |
| 208 | 420 | 13.50(0.16) | 13.65(0.26) | 14.34(0.35) | 14.16(0.32) | 14.45(3.00) | 13.72(0.10) | 13.71(0.11) |
| 208 | 264 | 13.26(0.16) | 13.28(0.15) | 13.67(0.27) | 13.49(0.22) | 14.98(1.64) | 13.40(0.10) | 13.41(0.10) |
| 208 | 243 | 13.27(0.17) | 13.29(0.18) | 13.82(0.30) | 13.60(0.23) | 14.87(1.61) | 13.44(0.08) | 13.41(0.09) |
| 208 | 338 | 13.27(0.15) | 13.25(0.14) | 13.59(0.27) | 13.46(0.21) | 14.76(2.79) | 13.49(0.09) | 13.50(0.10) |
| 208 | 73 | 13.49(0.15) | 13.51(0.17) | 14.00(0.31) | 13.81(0.24) | 14.91(2.60) | 13.76(0.12) | 13.73(0.12) |
| 208 | 458 | 13.53(0.24) | 13.51(0.24) | 13.89(0.33) | 13.73(0.26) | 15.10(2.32) | 13.77(0.23) | 13.77(0.22) |
| 208 | 167 | 13.39(0.14) | 13.39(0.15) | 13.79(0.25) | 13.59(0.19) | 14.92(1.01) | 13.65(0.11) | 13.69(0.11) |
| 208 | 443 | 13.59(0.13) | 13.58(0.14) | 13.87(0.22) | 13.76(0.18) | 14.90(1.73) | 13.87(0.12) | 13.87(0.10) |
| 208 | 300 | 13.35(0.16) | 13.30(0.16) | 13.65(0.22) | 13.54(0.19) | 15.01(1.95) | 13.55(0.12) | 13.54(0.12) |
| 300 | 420 | 13.48(0.16) | 13.64(0.32) | 14.21(0.28) | 14.08(0.27) | 14.15(1.46) | 13.71(0.09) | 13.71(0.10) |
| 300 | 264 | 13.19(0.13) | 13.17(0.18) | 13.51(0.23) | 13.40(0.20) | 14.35(1.58) | 13.41(0.10) | 13.43(0.09) |
| 300 | 243 | 13.21(0.16) | 13.25(0.22) | 13.64(0.26) | 13.53(0.25) | 14.15(2.45) | 13.41(0.10) | 13.43(0.10) |
| 300 | 338 | 13.21(0.13) | 13.19(0.14) | 13.48(0.20) | 13.41(0.19) | 14.26(2.29) | 13.49(0.09) | 13.51(0.09) |
| 300 | 73 | 13.46(0.15) | 13.46(0.26) | 13.85(0.21) | 13.70(0.20) | 14.33(1.92) | 13.74(0.12) | 13.73(0.12) |
| 300 | 458 | 13.46(0.26) | 13.46(0.28) | 13.75(0.27) | 13.64(0.25) | 14.40(1.37) | 13.76(0.18) | 13.76(0.22) |
| 300 | 167 | 13.37(0.14) | 13.36(0.20) | 13.66(0.24) | 13.56(0.20) | 14.38(1.77) | 13.66(0.11) | 13.65(0.10) |
| 300 | 443 | 13.58(0.14) | 13.56(0.17) | 13.79(0.16) | 13.73(0.15) | 14.53(1.26) | 13.88(0.12) | 13.89(0.12) |
| 300 | 208 | 13.33(0.17) | 13.32(0.22) | 13.62(0.22) | 13.52(0.21) | 14.49(2.42) | 13.64(0.12) | 13.67(0.12) |
| Population Median |  | 13.36(0.24) | 13.38(0.26) | 13.68(0.37) | 13.58(0.35) | 15.30(2.37) | 13.62(0.19) | 13.62(0.19) |
| Outperformance ratio of *OTTEHR* |  | N.A. | 1.98 | 27.39 | 19.75 | 89.87 | 22.89 | 22.90 |

**Table S10:** Benchmark results for pairwise cross-hospital experiments. Medians and standard deviations of the log of *RMSE* between projected duration and observed duration on target admissions for different insurance groups using *OTTEHR, TCA, CA, GFK, deepJDOT, RSD* and *daregram*. The outperformance ratio of *OTTEHR* to *TCA*/*CA*/*GFK*/*RSD*/*daregram* is defined as the percentage decrease in the median *RMSE* from *TCA*/*CA*/*GFK*/*RSD*/*daregram* to *OTTEHR*. The outperformance ratio of *OTTEHR* to *deepJDOT* is defined as the percentage decrease in median of the log-transformed of standard deviation of *RMSE* from *deepJDOT* to *OTTEHR*.

| Source | Target | *OTTEHR* | *TCA* | *CA* | *GFK* | *deepJDOT* | *RSD* | *daregram* |
| --- | --- | --- | --- | --- | --- | --- | --- | --- |
| 420 | 264 | 13.46(0.17) | 13.43(0.15) | 13.54(0.16) | 13.52(0.15) | 19.02(1.90) | 13.70(0.12) | 13.74(0.14) |
| 420 | 243 | 13.46(0.21) | 13.45(0.17) | 13.64(0.17) | 13.54(0.17) | 18.80(1.92) | 13.74(0.16) | 13.74(0.16) |
| 420 | 338 | 13.49(0.16) | 13.46(0.15) | 13.57(0.14) | 13.54(0.14) | 18.60(1.72) | 13.78(0.12) | 13.81(0.13) |
| 420 | 73 | 13.88(0.26) | 13.79(0.26) | 13.91(0.23) | 13.85(0.24) | 18.64(1.76) | 14.14(0.19) | 14.13(0.18) |
| 420 | 458 | 14.03(0.69) | 13.97(0.71) | 14.03(0.67) | 14.00(0.69) | 18.71(1.34) | 14.28(0.51) | 14.22(0.54) |
| 420 | 167 | 13.73(0.22) | 13.65(0.22) | 13.80(0.20) | 13.71(0.21) | 18.43(1.33) | 14.02(0.17) | 14.00(0.16) |
| 420 | 443 | 14.08(0.17) | 14.01(0.17) | 14.04(0.16) | 14.03(0.16) | 18.87(1.49) | 14.29(0.15) | 14.27(0.15) |
| 420 | 208 | 13.80(0.30) | 13.72(0.30) | 13.80(0.28) | 13.78(0.29) | 18.80(1.97) | 14.01(0.24) | 14.07(0.28) |
| 420 | 300 | 13.63(0.26) | 13.60(0.25) | 13.68(0.23) | 13.64(0.24) | 18.91(1.80) | 13.93(0.22) | 13.88(0.19) |
| 264 | 420 | 13.87(0.17) | 13.95(0.19) | 14.31(0.18) | 14.17(0.14) | 14.11(1.90) | 14.00(0.14) | 14.05(0.17) |
| 264 | 243 | 13.49(0.19) | 13.53(0.21) | 13.82(0.20) | 13.71(0.18) | 14.20(1.78) | 13.74(0.16) | 13.73(0.16) |
| 264 | 338 | 13.51(0.17) | 13.47(0.17) | 13.66(0.18) | 13.60(0.17) | 14.31(1.29) | 13.81(0.13) | 13.79(0.12) |
| 264 | 73 | 13.92(0.23) | 13.90(0.23) | 14.11(0.19) | 14.01(0.19) | 14.49(1.08) | 14.15(0.18) | 14.14(0.19) |
| 264 | 458 | 14.02(0.62) | 14.02(0.62) | 14.11(0.57) | 14.10(0.59) | 14.52(2.00) | 14.20(0.57) | 14.14(0.41) |
| 264 | 167 | 13.75(0.20) | 13.75(0.20) | 13.93(0.19) | 13.84(0.18) | 14.61(2.87) | 14.05(0.15) | 14.00(0.16) |
| 264 | 443 | 14.09(0.19) | 14.06(0.19) | 14.16(0.17) | 14.12(0.17) | 14.45(1.63) | 14.28(0.15) | 14.29(0.15) |
| 264 | 208 | 13.77(0.33) | 13.76(0.33) | 13.90(0.29) | 13.83(0.30) | 14.47(1.20) | 14.02(0.27) | 14.04(0.25) |
| 264 | 300 | 13.66(0.26) | 13.64(0.27) | 13.80(0.22) | 13.73(0.23) | 14.34(1.92) | 13.88(0.19) | 13.91(0.20) |
| 243 | 420 | 13.83(0.19) | 13.85(0.21) | 14.19(0.21) | 14.09(0.18) | 14.81(1.88) | 14.05(0.15) | 14.05(0.17) |
| 243 | 264 | 13.45(0.17) | 13.38(0.18) | 13.62(0.17) | 13.56(0.16) | 15.08(1.35) | 13.70(0.15) | 13.73(0.16) |
| 243 | 338 | 13.45(0.18) | 13.44(0.17) | 13.59(0.18) | 13.57(0.17) | 15.04(1.97) | 13.77(0.13) | 13.84(0.14) |
| 243 | 73 | 13.94(0.26) | 13.90(0.27) | 14.07(0.22) | 14.00(0.23) | 15.17(1.83) | 14.14(0.19) | 14.12(0.19) |
| 243 | 458 | 14.05(0.74) | 14.02(0.75) | 14.13(0.70) | 14.06(0.72) | 15.43(2.55) | 14.21(0.47) | 14.27(0.46) |
| 243 | 167 | 13.75(0.20) | 13.73(0.20) | 13.90(0.18) | 13.76(0.19) | 15.22(1.52) | 14.01(0.18) | 14.03(0.15) |
| 243 | 443 | 14.07(0.19) | 14.04(0.20) | 14.10(0.17) | 14.08(0.17) | 15.12(1.63) | 14.27(0.14) | 14.31(0.16) |
| 243 | 208 | 13.80(0.32) | 13.76(0.33) | 13.87(0.29) | 13.83(0.30) | 15.13(1.64) | 14.04(0.23) | 14.05(0.24) |
| 243 | 300 | 13.64(0.30) | 13.61(0.30) | 13.74(0.26) | 13.68(0.27) | 15.11(1.07) | 13.90(0.21) | 13.91(0.21) |
| 338 | 420 | 13.82(0.19) | 13.95(0.17) | 14.40(0.21) | 14.25(0.14) | 14.15(1.59) | 14.04(0.15) | 14.04(0.14) |
| 338 | 264 | 13.47(0.16) | 13.46(0.16) | 13.72(0.17) | 13.60(0.13) | 14.34(2.27) | 13.74(0.13) | 13.73(0.14) |
| 338 | 243 | 13.50(0.19) | 13.55(0.19) | 13.86(0.20) | 13.76(0.17) | 14.04(1.22) | 13.76(0.15) | 13.72(0.14) |
| 338 | 73 | 13.89(0.21) | 13.93(0.22) | 14.16(0.20) | 14.03(0.18) | 14.46(1.87) | 14.13(0.18) | 14.11(0.19) |
| 338 | 458 | 14.06(0.63) | 14.05(0.63) | 14.17(0.58) | 14.10(0.60) | 14.57(2.03) | 14.24(0.53) | 14.24(0.35) |
| 338 | 167 | 13.72(0.20) | 13.74(0.19) | 13.95(0.20) | 13.84(0.18) | 14.43(1.81) | 14.03(0.17) | 14.03(0.15) |
| 338 | 443 | 14.07(0.20) | 14.05(0.20) | 14.18(0.19) | 14.12(0.19) | 14.51(1.74) | 14.27(0.15) | 14.31(0.13) |
| 338 | 208 | 13.76(0.26) | 13.76(0.26) | 13.95(0.23) | 13.85(0.24) | 14.44(1.05) | 14.05(0.23) | 14.05(0.26) |
| 338 | 300 | 13.61(0.26) | 13.62(0.27) | 13.81(0.23) | 13.72(0.24) | 14.30(2.50) | 13.89(0.20) | 13.92(0.21) |
| 73 | 420 | 13.91(0.18) | 13.94(0.18) | 14.39(0.19) | 14.24(0.19) | 16.32(2.32) | 14.05(0.13) | 14.06(0.16) |
| 73 | 264 | 13.47(0.16) | 13.52(0.14) | 13.77(0.16) | 13.69(0.17) | 16.84(2.09) | 13.72(0.12) | 13.70(0.15) |
| 73 | 243 | 13.63(0.21) | 13.60(0.19) | 13.97(0.20) | 13.82(0.18) | 16.30(2.31) | 13.77(0.15) | 13.75(0.14) |
| 73 | 338 | 13.49(0.17) | 13.52(0.15) | 13.79(0.18) | 13.72(0.17) | 16.67(1.91) | 13.80(0.14) | 13.79(0.13) |
| 73 | 458 | 14.04(0.64) | 14.00(0.64) | 14.20(0.57) | 14.12(0.59) | 16.49(2.87) | 14.27(0.50) | 14.19(0.55) |
| 73 | 167 | 13.78(0.19) | 13.75(0.19) | 13.97(0.19) | 13.88(0.18) | 16.60(1.89) | 14.03(0.14) | 14.03(0.16) |
| 73 | 443 | 14.06(0.19) | 14.03(0.19) | 14.18(0.17) | 14.15(0.18) | 16.51(1.97) | 14.30(0.13) | 14.26(0.13) |
| 73 | 208 | 13.79(0.29) | 13.77(0.29) | 14.00(0.25) | 13.91(0.25) | 17.01(2.07) | 14.05(0.22) | 14.02(0.24) |
| 73 | 300 | 13.64(0.27) | 13.66(0.25) | 13.86(0.20) | 13.79(0.22) | 16.88(1.74) | 13.88(0.22) | 13.89(0.22) |
| 458 | 420 | 14.07(0.68) | 14.10(0.53) | 14.73(0.66) | 14.54(0.70) | 15.41(2.79) | 14.06(0.14) | 14.04(0.14) |
| 458 | 264 | 13.67(0.33) | 13.62(0.34) | 14.03(0.53) | 13.93(0.55) | 16.31(3.35) | 13.75(0.16) | 13.71(0.14) |
| 458 | 243 | 13.76(0.53) | 13.69(0.51) | 14.24(0.59) | 14.05(0.64) | 15.90(1.92) | 13.75(0.12) | 13.73(0.16) |
| 458 | 338 | 13.65(0.31) | 13.59(0.40) | 14.04(0.51) | 13.93(0.54) | 15.67(1.99) | 13.80(0.12) | 13.78(0.12) |
| 458 | 73 | 14.06(0.55) | 13.98(0.42) | 14.42(0.57) | 14.24(0.57) | 15.69(2.66) | 14.15(0.19) | 14.19(0.18) |
| 458 | 167 | 13.85(0.41) | 13.86(0.31) | 14.19(0.48) | 14.02(0.52) | 15.89(1.65) | 13.99(0.16) | 14.00(0.14) |
| 458 | 443 | 14.15(0.25) | 14.12(0.22) | 14.37(0.44) | 14.28(0.46) | 15.83(2.04) | 14.27(0.13) | 14.30(0.14) |
| 458 | 208 | 13.87(0.38) | 13.81(0.35) | 14.19(0.47) | 14.01(0.51) | 16.04(1.99) | 14.03(0.24) | 14.03(0.22) |
| 458 | 300 | 13.81(0.31) | 13.76(0.33) | 14.22(0.49) | 14.08(0.51) | 15.66(2.29) | 13.90(0.23) | 13.87(0.22) |

**Table S11:** Benchmark results for pairwise cross-hospital experiments (continued). Medians and standard deviations of the log of *RMSE* between projected duration and observed duration on target admissions for different insurance groups using *OTTEHR, TCA, CA, GFK, deepJDOT, RSD* and *daregram*. The outperformance ratio of *OTTEHR* to *TCA*/*CA*/*GFK*/*RSD*/*daregram* is defined as the percentage decrease in the median *RMSE* from *TCA*/*CA*/*GFK*/*RSD*/*daregram* to *OTTEHR*. The outperformance ratio of *OTTEHR* to *deepJDOT* is defined as the percentage decrease in median of the log-transformed of standard deviation of *RMSE* from *deepJDOT* to *OTTEHR*.

| Source | Target | *OTTEHR* | *TCA* | *CA* | *GFK* | *deepJDOT* | *RSD* | *daregram* |
| --- | --- | --- | --- | --- | --- | --- | --- | --- |
| 167 | 420 | 13.93(0.19) | 13.98(0.28) | 14.45(0.22) | 14.28(0.20) | 15.05(2.10) | 14.08(0.17) | 14.04(0.14) |
| 167 | 264 | 13.54(0.14) | 13.53(0.18) | 13.80(0.16) | 13.71(0.14) | 15.84(2.27) | 13.72(0.14) | 13.73(0.14) |
| 167 | 243 | 13.57(0.17) | 13.54(0.19) | 13.97(0.20) | 13.83(0.17) | 15.36(1.71) | 13.77(0.15) | 13.77(0.15) |
| 167 | 338 | 13.57(0.16) | 13.52(0.15) | 13.77(0.18) | 13.68(0.15) | 15.54(2.77) | 13.79(0.15) | 13.79(0.11) |
| 167 | 73 | 13.86(0.20) | 13.87(0.21) | 14.18(0.18) | 14.05(0.19) | 15.59(1.38) | 14.14(0.19) | 14.12(0.21) |
| 167 | 458 | 14.01(0.63) | 14.00(0.64) | 14.17(0.56) | 14.08(0.60) | 15.77(2.76) | 14.28(0.56) | 14.28(0.52) |
| 167 | 443 | 14.05(0.20) | 14.01(0.19) | 14.17(0.16) | 14.12(0.18) | 15.52(2.52) | 14.29(0.16) | 14.28(0.18) |
| 167 | 208 | 13.75(0.30) | 13.74(0.30) | 13.97(0.26) | 13.85(0.27) | 15.78(1.72) | 14.06(0.23) | 14.08(0.25) |
| 167 | 300 | 13.72(0.26) | 13.67(0.26) | 13.89(0.22) | 13.81(0.23) | 15.65(1.72) | 13.89(0.26) | 13.88(0.24) |
| 443 | 420 | 14.07(0.19) | 14.10(0.20) | 14.75(0.22) | 14.61(0.20) | 14.88(1.19) | 14.05(0.17) | 14.08(0.14) |
| 443 | 264 | 13.70(0.15) | 13.69(0.16) | 14.07(0.17) | 13.94(0.16) | 15.64(1.73) | 13.74(0.16) | 13.71(0.16) |
| 443 | 243 | 13.80(0.17) | 13.79(0.19) | 14.25(0.22) | 14.08(0.18) | 15.75(3.04) | 13.75(0.16) | 13.76(0.14) |
| 443 | 338 | 13.68(0.17) | 13.67(0.18) | 14.01(0.18) | 13.92(0.18) | 15.79(1.29) | 13.80(0.12) | 13.79(0.12) |
| 443 | 73 | 13.98(0.19) | 14.01(0.21) | 14.38(0.21) | 14.27(0.20) | 15.87(2.35) | 14.14(0.21) | 14.13(0.21) |
| 443 | 458 | 14.12(0.61) | 14.13(0.62) | 14.38(0.52) | 14.27(0.54) | 15.77(1.03) | 14.27(0.47) | 14.28(0.51) |
| 443 | 167 | 13.84(0.18) | 13.88(0.20) | 14.21(0.21) | 14.05(0.19) | 15.66(1.46) | 14.04(0.16) | 14.03(0.17) |
| 443 | 208 | 13.87(0.26) | 13.85(0.26) | 14.14(0.21) | 14.07(0.22) | 15.90(1.15) | 14.05(0.22) | 14.06(0.23) |
| 443 | 300 | 13.76(0.21) | 13.74(0.22) | 14.06(0.19) | 14.00(0.18) | 15.72(2.28) | 13.89(0.20) | 13.89(0.21) |
| 208 | 420 | 13.93(0.20) | 14.04(0.27) | 14.62(0.34) | 14.45(0.30) | 14.61(2.92) | 14.09(0.13) | 14.05(0.17) |
| 208 | 264 | 13.60(0.21) | 13.60(0.20) | 13.90(0.27) | 13.77(0.22) | 15.00(1.55) | 13.69(0.15) | 13.73(0.15) |
| 208 | 243 | 13.64(0.21) | 13.63(0.23) | 14.12(0.29) | 13.91(0.23) | 14.89(1.48) | 13.77(0.13) | 13.74(0.14) |
| 208 | 338 | 13.60(0.21) | 13.55(0.19) | 13.86(0.27) | 13.75(0.22) | 14.81(2.73) | 13.79(0.13) | 13.81(0.12) |
| 208 | 73 | 13.94(0.22) | 13.98(0.22) | 14.34(0.31) | 14.18(0.25) | 15.05(2.52) | 14.13(0.21) | 14.13(0.18) |
| 208 | 458 | 14.07(0.54) | 14.07(0.55) | 14.30(0.51) | 14.18(0.50) | 15.25(2.22) | 14.27(0.58) | 14.27(0.51) |
| 208 | 167 | 13.81(0.19) | 13.81(0.21) | 14.09(0.24) | 13.94(0.20) | 14.99(0.90) | 14.03(0.15) | 14.05(0.16) |
| 208 | 443 | 14.09(0.17) | 14.08(0.19) | 14.26(0.20) | 14.17(0.18) | 15.01(1.65) | 14.29(0.15) | 14.30(0.12) |
| 208 | 300 | 13.76(0.27) | 13.67(0.28) | 14.01(0.26) | 13.89(0.25) | 15.03(1.87) | 13.90(0.22) | 13.91(0.21) |
| 300 | 420 | 13.92(0.21) | 14.01(0.30) | 14.50(0.27) | 14.35(0.25) | 14.34(1.35) | 14.07(0.14) | 14.07(0.14) |
| 300 | 264 | 13.53(0.18) | 13.50(0.24) | 13.81(0.23) | 13.71(0.21) | 14.44(1.49) | 13.74(0.15) | 13.76(0.15) |
| 300 | 243 | 13.58(0.21) | 13.61(0.30) | 13.95(0.24) | 13.83(0.24) | 14.29(2.35) | 13.75(0.14) | 13.74(0.14) |
| 300 | 338 | 13.58(0.17) | 13.52(0.21) | 13.76(0.19) | 13.70(0.19) | 14.38(2.20) | 13.79(0.13) | 13.83(0.13) |
| 300 | 73 | 13.93(0.23) | 13.99(0.28) | 14.23(0.21) | 14.12(0.21) | 14.46(1.84) | 14.15(0.20) | 14.10(0.19) |
| 300 | 458 | 14.08(0.59) | 14.09(0.61) | 14.20(0.54) | 14.16(0.55) | 14.73(1.23) | 14.24(0.49) | 14.24(0.55) |
| 300 | 167 | 13.79(0.20) | 13.77(0.24) | 14.03(0.23) | 13.93(0.21) | 14.51(1.68) | 14.03(0.17) | 14.02(0.15) |
| 300 | 443 | 14.10(0.17) | 14.08(0.20) | 14.21(0.17) | 14.17(0.17) | 14.72(1.14) | 14.30(0.16) | 14.32(0.15) |
| 300 | 208 | 13.76(0.31) | 13.75(0.34) | 13.97(0.28) | 13.88(0.29) | 14.68(2.33) | 14.02(0.22) | 14.08(0.25) |
| Population Median |  | 13.79(0.38) | 13.78(0.38) | 14.05(0.42) | 13.95(0.41) | 15.34(2.28) | 13.99(0.31) | 13.99(0.31) |
| Outperformance ratio of *OTTEHR* |  | N.A. | -1.00 | 22.90 | 14.79 | 83.33 | 18.13 | 18.13 |

### E.3 Computation time

**Table S12:**
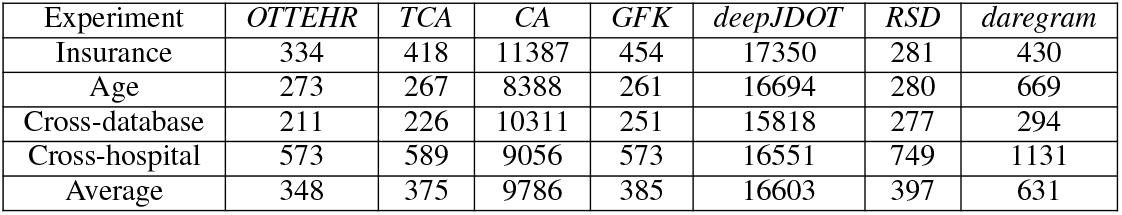
Average computational time in seconds per experiment for *OTTEHR, TCA, CA, GFK, deepJDOT, RSD* and *daregram*.

| Experiment | *OTTEHR* | *TCA* | *CA* | *GFK* | *deepJDOT* | *RSD* | *daregram* |
| --- | --- | --- | --- | --- | --- | --- | --- |
| Insurance | 334 | 418 | 11387 | 454 | 17350 | 281 | 430 |
| Age | 273 | 267 | 8388 | 261 | 16694 | 280 | 669 |
| Cross-database | 211 | 226 | 10311 | 251 | 15818 | 277 | 294 |
| Cross-hospital | 573 | 589 | 9056 | 573 | 16551 | 749 | 1131 |
| Average | 348 | 375 | 9786 | 385 | 16603 | 397 | 631 |

## Notes

### Competing Interest Statement

The authors have declared no competing interest.

### Funding Statement

This study did not receive any external funding.

### Author Declarations

The study used only openly available human data that were originally located at Physionet.

### Summary of Updates

We added more experiments on MIMIC-III, MIMIC-IV and eICU dataset.

